# Inferring causal pathways between metabolic processes and liver fat accumulation: an IMI DIRECT study

**DOI:** 10.1101/2021.08.31.21262709

**Authors:** Naeimeh Atabaki-Pasdar, Hugo Pomares-Millan, Robert W Koivula, Andrea Tura, Andrew Brown, Ana Viñuela, Leandro Agudelo, Daniel Coral, Sabine van Oort, Kristine Allin, Elizaveta Chabanova, Henna Cederberg, Federico De Masi, Petra Elders, Juan Fernandez Tajes, Ian M Forgie, Tue H Hansen, Alison Heggie, Angus Jones, Tarja Kokkola, Anubha Mahajan, Timothy J McDonald, Donna McEvoy, Konstantinos Tsirigos, Harriet Teare, Jagadish Vangipurapu, Henrik Vestergaard, Jerzy Adamski, Joline WJ Beulens, Søren Brunak, Emmanouil Dermitzakis, Torben Hansen, Andrew T Hattersley, Markku Laakso, Oluf Pedersen, Martin Ridderstråle, Hartmut Ruetten, Femke Rutters, Jochen M Schwenk, Mark Walker, Giuseppe N Giordano, Mattias Ohlsson, Ramneek Gupta, Andrea Mari, Mark I McCarthy, E Louise Thomas, Jimmy D Bell, Imre Pavo, Ewan R Pearson, Paul W Franks

## Abstract

Type 2 diabetes (T2D) and non-alcoholic fatty liver disease (NAFLD) often co-occur. Defining causal pathways underlying this relationship may help optimize the prevention and treatment of both diseases. Thus, we assessed the strength and magnitude of the putative causal pathways linking dysglycemia and fatty liver, using a combination of causal inference methods.

Measures of glycemia, insulin dynamics, magnetic resonance imaging (MRI)-derived abdominal and liver fat content, serological biomarkers, lifestyle, and anthropometry were obtained in participants from the IMI DIRECT cohorts (n=795 with new onset T2D and 2234 individuals free from diabetes). UK Biobank (n=3641) was used for modelling and replication purposes. Bayesian networks were employed to infer causal pathways, with causal validation using two-sample Mendelian randomization.

Bayesian networks fitted to IMI DIRECT data identified higher basal insulin secretion rate (BasalISR) and MRI-derived excess visceral fat (VAT) accumulation as the features of dysmetabolism most likely to cause liver fat accumulation; the unconditional probability of fatty liver (>5%) increased significantly when conditioning on high levels of BasalISR and VAT (by 23%, 32% respectively; 40% for both). Analyses in UK Biobank yielded comparable results. MR confirmed most causal pathways predicted by the Bayesian networks.

Here, BasalISR had the highest causal effect on fatty liver predisposition, providing mechanistic evidence underpinning the established association of NAFLD and T2D. BasalISR may represent a pragmatic biomarker for NAFLD prediction in clinical practice.

## INTRODUCTION

Around 20-30% of adults in high-income countries have non-alcoholic fatty liver disease (NAFLD), with rates increasing worldwide^1^. For several decades, excessive intrahepatic fat accumulation has been recognized as a risk factor for type 2 diabetes (T2D)^2^, yet the evidence for this stem largely from cross-sectional studies^3^, which are prone to bias, confounding and reverse causation. Determining the causal nature of these relationships and their underlying metabolic mechanisms is necessary to optimize the prevention and treatment of NAFLD and T2D.

NAFLD is highly heritable (h^2^∼ 40%)^4^, suggesting that, as with T2D, inherited DNA variants affect predisposition. Nevertheless, both diseases are also caused by numerous obesogenic environmental factors^5^. Excessive weight gain can cause fatty liver by initially raising circulating free fatty acid concentrations, which are then taken up by hepatocytes and stored as triacylglycerol. This in turn can trigger a cycle of liver-specific insulin resistance and excessive *de novo* hepatic glucose and lipid production^6-8^. This cycle of dyslipidemia and dysglycemia is postulated to link obesity, fatty liver and diabetes^9-11^.

Fatty liver pathology ranges from simple steatosis, through non-alcoholic steatohepatitis (NASH), liver fibrosis and ultimately cirrhosis. T2D and cardiovascular disease often co-occur with NAFLD, but the causal relationships are poorly defined^12^. Here, we examined a range of putative causal pathways linking the development of NAFLD with T2D using Bayesian network (BN) and bidirectional Mendelian randomization (MR) analyses.

## RESULTS

The primary analyses were conducted within IMI DIRECT, a multicenter prospective cohort study of 3029 European-ancestry adults diagnosed with T2D (n=795) or without T2D (n=2234); the latter group included participants with normal and prediabetic glycemia (ascertained by HbA1c, fasting glucose or 2-hour glucose). Of these participants, 1070 had the required variables for a complete case analysis with the mean (SD) age of 62 (5.0) years and 77.5% being male. In addition, the UK Biobank was utilized with the analysis sample restricted to white European ethnicity (n= 442580) and complete cases (n= 3641); in UK Biobank, the mean (SD) age of the cohort was 56.8 (8.0) years and 47.2% were male.

The following section describes the BNs in the IMI DIRECT and UK Biobank cohorts (subgroup analyses were performed in participants with and without T2D and in both sexes). The bidirectional two-sample MR results that were statistically significant after Benjamini-Hochberg False Discovery Rate (FDR) correction, at 5% threshold^13^ per trait, were investigated for the directed association (arcs) that were present in the BNs. Detailed results for all bidirectional MR analyses performed here, including information about the published genome-wide association studies (GWAS) from which the genetic instruments are derived, are reported separately and per trait (S1 Table). The subset of statistically significant MR results is summarized in Table 1.

**Table 1.** Two-sample MR exposure-outcome significant associations that achieved corrected threshold and sensitivity analysis.

|  |  |  | IVW |  |  |  |  | MR-Egger |  |  |  | Sensitivity analysis |  |  |
| --- | --- | --- | --- | --- | --- | --- | --- | --- | --- | --- | --- | --- | --- | --- |
| Exposure | Outcome | # SNPs | $\beta$ | $\beta SE$ | P-value | P-value<br>FDR corrected | | $\beta$ | $\beta SE$ | P-value | | MR-Egger<br>Intercept | MR-Egger<br>Intercept P-value | MR-PRESSO<br>global test P-value |
| Glycemic traits |  |  |  |  |  |  |  |  |  |  |  |  |  |  |
| HbA1C | HDL | 11 | 0.107 | 0.037 | 0.004 | 0.010 |  | 0.205 | 0.091 | 0.050 |  | -0.004 | 0.236 | 0.543 |
| HbA1C | ISI | 11 | -0.317 | 0.118 | 0.007 | 0.019 |  | -0.291 | 0.321 | 0.389 |  | -0.001 | 0.931 | 0.830 |
| Adiposity/obesity |  |  |  |  |  |  |  |  |  |  |  |  |  |  |
| BMI | Serum albumin level | 62 | -0.573 | 0.221 | 0.010 | 0.024 |  | -0.873 | 0.638 | 0.177 |  | 0.017 | 0.232 | 0.046 |
| BMI | Fasting blood insulin | 67 | 0.172 | 0.019 | 6.25E-20 | 3.92E-19 |  | 0.126 | 0.055 | 0.025 |  | 0.001 | 0.464 | 0.073 |
| BMI | Fasting blood glucose | 66 | 0.058 | 0.024 | 0.016 | 0.039 |  | 0.109 | 0.070 | 0.124 |  | -0.001 | 0.672 | 0.157 |
| Waist circumference | Serum albumin levels | 35 | -0.678 | 0.272 | 0.013 | 0.032 |  | -0.701 | 0.934 | 0.458 |  | 0.014 | 0.437 | 0.875 |
| Waist circumference | Fasting blood insulin | 41 | 0.232 | 0.025 | 1.11E-20 | 7.02E-20 |  | 0.141 | 0.087 | 0.114 |  | 0.002 | 0.19 | 0.064 |
| Whole body fat mass | Modified Stumvoll Insulin Sensitivity Index (model adjusted for BMI) | 184 | 0.111 | 0.045 | 0.013 | 0.032 |  | 0.133 | 0.137 | 0.333 |  | 0.0002 | 0.877 | 0.078 |
| Blood pressure |  |  |  |  |  |  |  |  |  |  |  |  |  |  |
| SBP | Two-hour glucose challenge | 108 | 0.271 | 0.108 | 0.012 | 0.030 |  | 0.462 | 0.416 | 0.269 |  | -0.004 | 0.615 | 0.497 |

We considered as significant if the directions of the estimates by IVW and MR-Egger were consistent, IVW method passed the FDR corrected threshold ( $p < 0.05$ ), and no significant pleiotropy tested by the MR-Egger intercept ( $p > 0.05$ ). Outlier SNPs were removed by MR-PRESSO at $p < 0.05$ global test. IVW: inverse variance weighted; MR-PRESSO: MR pleiotropy residual sum and outlier; ISI corresponds to Modified Stumvoll Insulin Sensitivity Index; BMI, body mass index; HbA1c, glycated hemoglobin A1C; HDL, fasting high-density lipoprotein cholesterol; SBP, systolic blood pressure; SNPs, single nucleotide polymorphisms.

### Bayesian Network and Mendelian randomization analyses in the IMI DIRECT cohorts

Prior to building the BNs, we applied cluster analysis to the input variables (the clustered variables are more likely to be present in the BN) (S1 Fig). A cluster comprising abdominal fat, body mass index (BMI), waist circumference and insulin measurements was observed in the Pearson correlation plot and the heatmap cluster analyses. These variables were initially selected based on their association with liver fat within the IMI DIRECT cohorts^14^ or from existing literature^15^. We then proceeded with structural and parameter learnings using the variables as nodes to build the BN. To have a more stable network with information on the strengths of the arcs amongst the nodes, we performed network averaging of the bootstrapped BN samples. Table 2 reports the conditional density and parameters for each node of the constructed BN. Fig 1A shows the averaged BN and Fig 1B shows the cumulative distribution function of the arc strengths. Detailed information of those arcs with strength and directional probabilities equal to or greater than the significant threshold (0.505, learned from the data) is reported in S2 Table along with the available MR results. Basal insulin secretion rate (BasalISR), visceral adipose tissue (VAT) and 2-hour insulin (TwoInsulin) were identified as the causal upstream (parent) nodes, whereas liver iron, alanine transaminase (ALT) and gamma-glutamyl transpeptidase (GGTP) were defined as the downstream (child) effects of liver fat, given the averaged BN estimates. *Markov blanket* of liver fat (the set of nodes that includes adequate knowledge for inference) was identified as liver iron, TwoInsulin, BasalISR, VAT, subcutaneous adipose tissue (SAT), BMI, glycated hemoglobin A1C (HbA1c), fasting glucose, oral glucose insulin sensitivity (OGIS)^16^, total fasting plasma GLP-1 (TotGLP1min0), mean insulin clearance (Clins), fasting triglycerides (TG), ALT, aspartate transaminase (AST) and GGTP (green nodes in Fig 1A). A more robust parsimonious network was generated by restricting the arcs to those with both strength and direction probabilities above or equal to 0.8 (Fig 1C); BasalISR, VAT and TwoInsulin remained as the causal nodes and liver iron as the downstream effect of liver fat. We then checked the averaged BN arcs with the two-sample MR results when available.

**Table 2.**
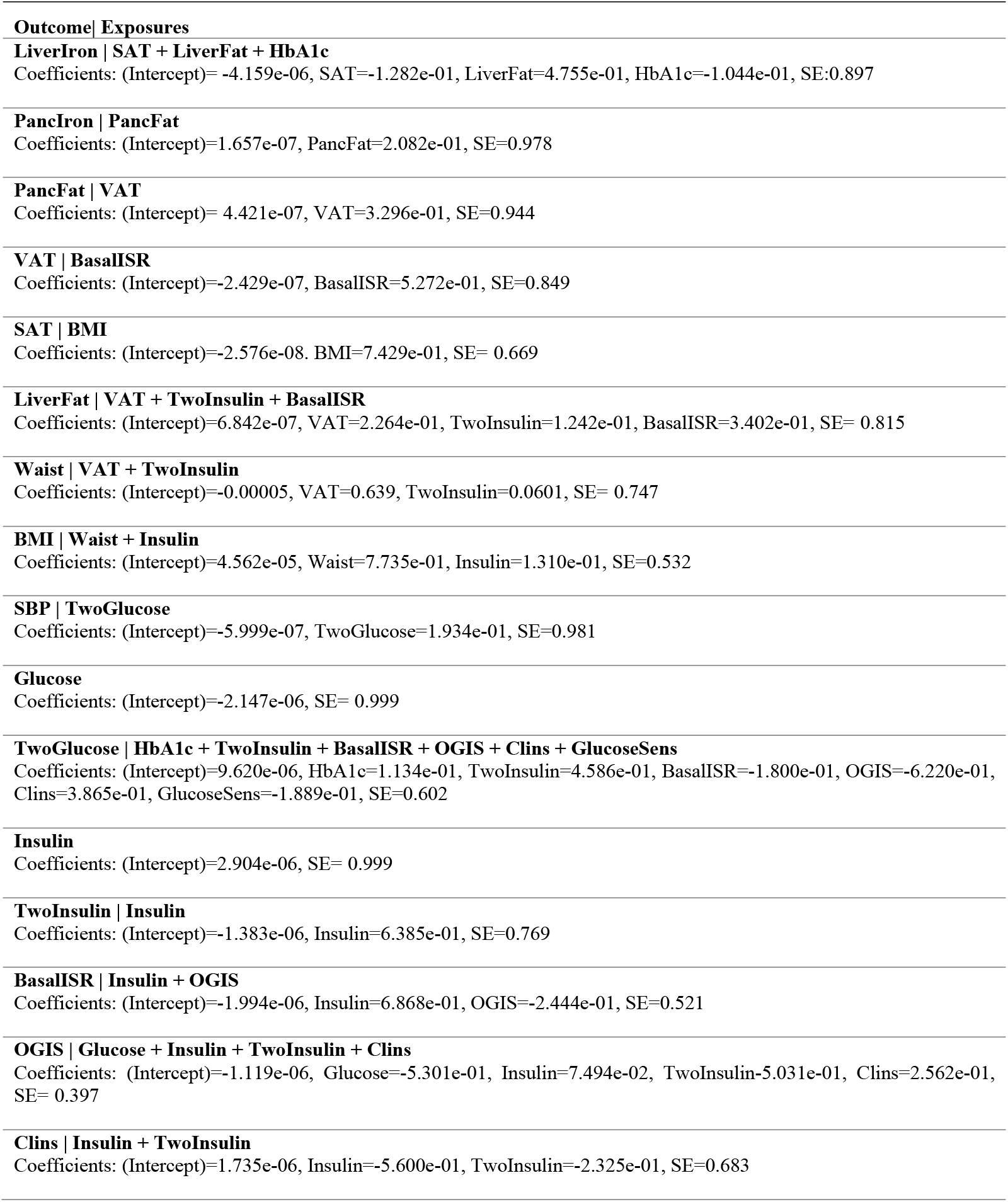
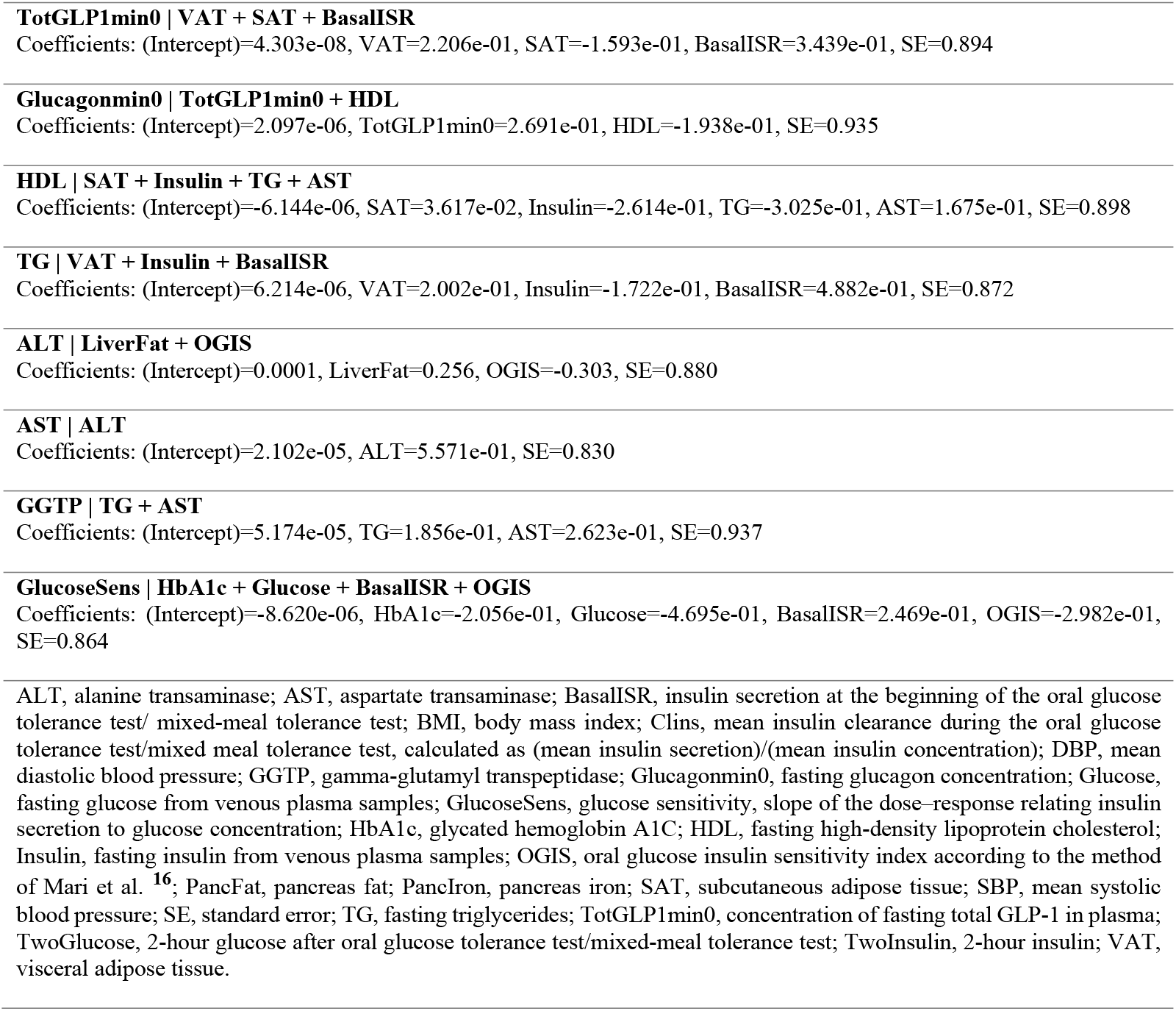
Conditional density and parameters for each node of the constructed average Bayesian network using IMI DIRECT inverse normal transformed data (n=1070).

**Fig 1.**
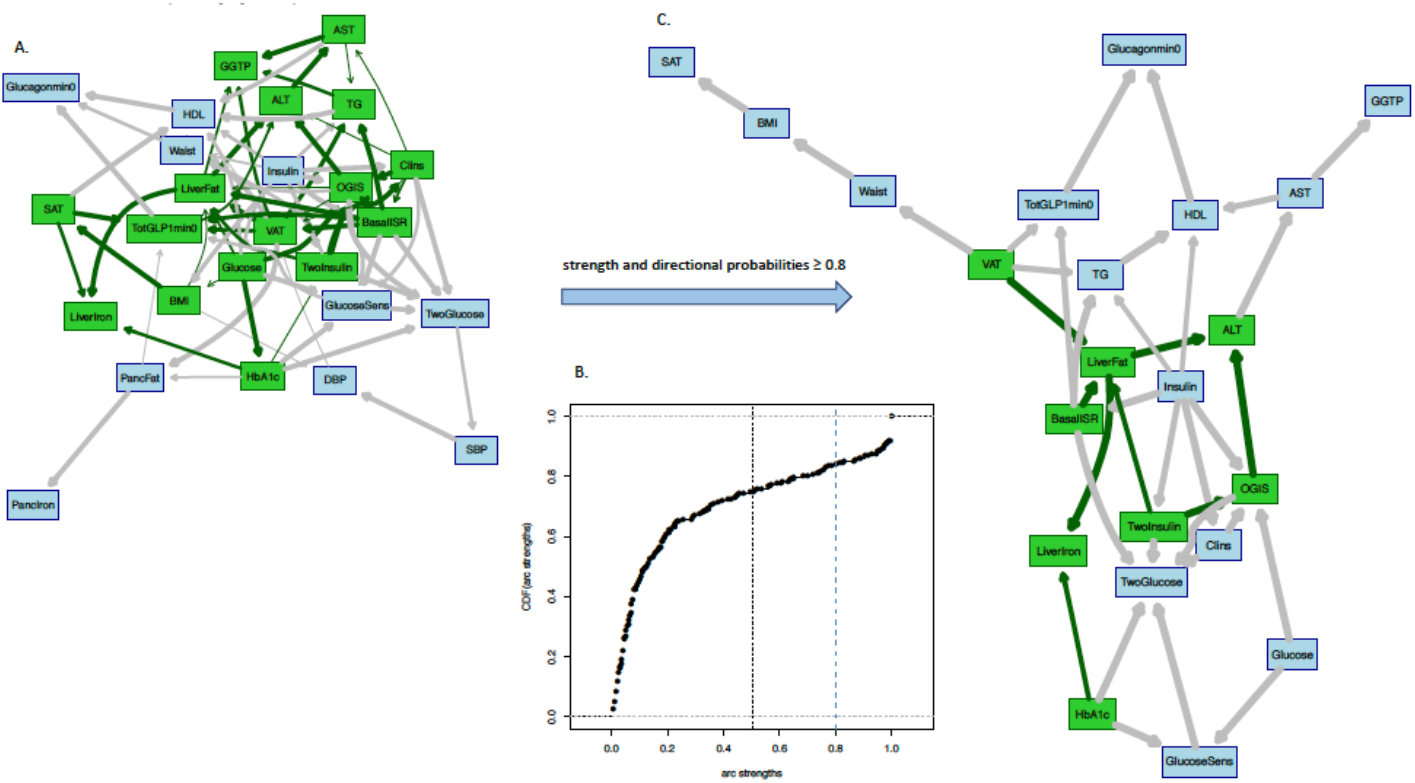
**Panel A:** Averaged Bayesian network of the bootstrapped samples among the variables of IMI DIRECT combined cohorts (data are inverse normal transformed, n=1070). **Panel B**: Cumulative distribution function of the arc strengths presented in the Bayesian network of Panel A. Black vertical line is the threshold (0.505) learned from data, equal or above which the arcs are identified as statistically significant. **Panel C**: Subset of Bayesian networks in Panel A including only arcs with strength and directional probabilities ≥ 0.8. Nodes in green highlight the Markov blanket of liver fat that includes the nodes with adequate information to stand as a separate Bayesian network. ALT, alanine transaminase; AST, aspartate transaminase; BasalISR, insulin secretion at the beginning of the oral glucose tolerance test/ mixed-meal tolerance test; BMI, body mass index; Clins, mean insulin clearance during the oral glucose tolerance test/mixed meal tolerance test, calculated as (mean insulin secretion)/(mean insulin concentration); DBP, mean diastolic blood pressure; GGTP, gamma-glutamyl transpeptidase; Glucagonmin0, fasting glucagon concentration; Glucose, fasting glucose from venous plasma samples; GlucoseSens, glucose sensitivity, slope of the dose–response relating insulin secretion to glucose concentration; HbA1c, glycated hemoglobin A1C; HDL, fasting high-density lipoprotein cholesterol; Insulin, fasting insulin from venous plasma samples; OGIS, oral glucose insulin sensitivity index according to the method of Mari et al. ^16^; PancFat, pancreas fat; PancIron, pancreas iron; SAT, subcutaneous adipose tissue; SBP, mean systolic blood pressure; TG, fasting triglycerides; TotGLP1min0, concentration of fasting total GLP-1 in plasma; TwoGlucose, 2-hour glucose after oral glucose tolerance test/mixed-meal tolerance test; TwoInsulin, 2-hour insulin; VAT, visceral adipose tissue.

In the MR analyses, several arcs revealed in the BN were consistent in directionality and magnitude (results shown in SD units, otherwise stated). Among these arcs, waist circumference had a likely causal link to BMI (β = 1.01; P_FDRcorrected_= 4.10E-198, P_Egger Intercept_ = 2.97E-01). Additionally, as in the BN, causal effects were inferred for SBP to DBP (β = 0.64; P_FDRcorrected_ = 5.95E-62, P_Egger Intercept_ = 5.2E-01). Moreover, AST (per lu/L increase) was causally associated with GGTP (lu/L) (β = 0.2; P_FDRcorrected_ = 1.14E-06, P_Egger Intercept_ = 1.52E-01) and waist circumference was inversely causally related with HDL (β = -0.31; P_FDRcorrected_ = 1.79E-10, P_Egger Intercept_ = 8.84E-01). A likely causal effect of TG on HDL (β = -0.12; P_FDRcorrected_ = 7.04E-04, P_Egger Intercept_ = 1.55E-01) was also observed.

### Subgroup analysis: diabetes vs. non-diabetes & female vs. male (in IMI DIRECT)

In order to elucidate causal networks that might be specific to diabetes or gender, analyses were reproduced within each subgroup. In the constructed BNs, BasalISR and VAT appeared to be the causal parental nodes for liver fat in both cohorts (S2 Fig, panels A and C). Subsets of these BNs with arcs having strengths and directional probabilities above or equal to 0.8 left only BasalISR as the causal parental node in both cohorts (S2 Fig, panels B and D). We then continued with the sensitivity analysis by gender, as illustrated in Figure S3. In the female’s BN, BasalISR and BMI were the causal parental nodes for liver fat, whereas in the male’s BN, BasalISR, VAT, TwoInsulin and HbA1c were the causal variables (S3 Fig, panel A and C). These BNs with extra restrictions on the strength and directional probabilities above or equal to 0.8 resulted in only BasalISR as the causal parental node for liver fat (S3 Fig, panel B and D). The Pearson correlation and the heatmap cluster analyses, performed in the sensitivity analyses resulted in similar clusters to the combined cohorts’ one (see S1, S4 and S5 Figs). Detailed information of arcs with strength and directional probabilities above or equal to the BNs’ thresholds (0.51, 0.5, 0.5 and 0.48 in the non-diabetes, diabetes, female and male BNs, respectively) are reported in S2 Table along with the corresponding MR results.

### Bayesian networks and posterior probabilistic inference (IMI DIRECT)

In order to calculate the posterior probabilities (updated probabilities given prior knowledge) of the conditional and unconditional queries, all continuous variables were first discretized into three levels (low, moderate and high) using Hartemink’s method^17^ where variables are transformed to discrete variables with he same number of levels (see S3 Table). Liver fat was considered as the variable of interest (*event*) where it was conditioned on some defined parental *evidence*; high BasalISR, high VAT and having high levels of both. Fig 2 summarizes unconditional and posterior conditional probabilities of liver fat on the combined, diabetes, non-diabetes, female and male groups of the IMI DIRECT BNs. A significant increase was observed after conditioning on high levels of BasalISR and VAT in the unconditional probability of high liver fat level on the random observations generated by the combined cohorts BNs: 23% increase after conditioning on VAT, 32% increase after conditioning on BasalISR, and 40% after conditioning on both. Similarly, diabetes, non-diabetes, female and male groups had substantial increments on the high liver fat level probability after conditioning on high levels of VAT, BasalISR and the combined effect. The probability of having high liver fat after conditioning on high levels of VAT and BasalISR was 0.84, 0.97, 0.86, 0.79 and 0.86 in the combined, diabetes, non-diabetes, female and male groups, respectively in the BNs constructed in IMI DIRECT.

**Fig 2.**
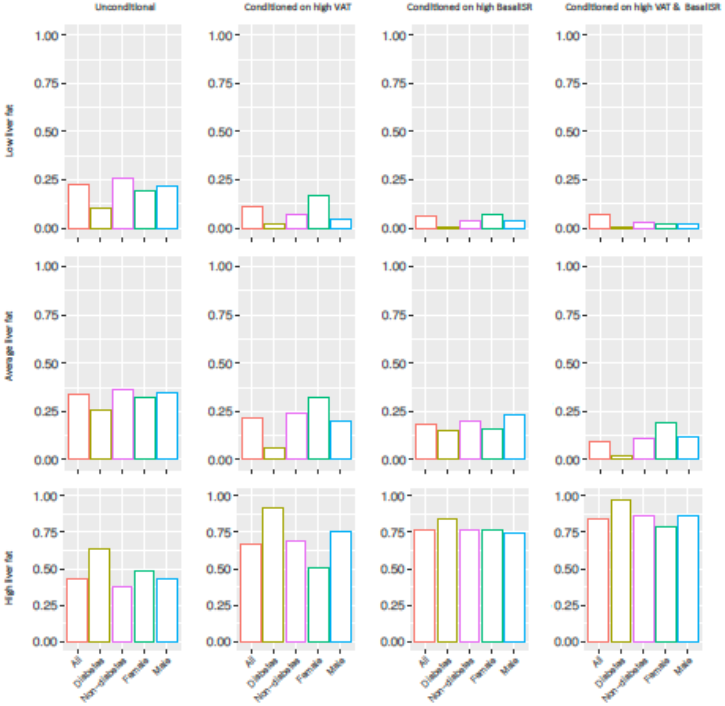
Unconditional and posterior conditional probabilities of liver fat on the combined (n=1070), diabetes (n=283), non-diabetes (n=787), female (n=239) and male (n=831) groups of the IMI DIRECT Bayesian networks (data are inverse normal transformed). Y axis represents the unconditional and posterior conditional probabilities of low (0.1,1.95], average (1.95,4.5] or high (4.5,37.6] levels of liver fat in rows. The posterior probabilities were obtained after conditioning on high levels of basal insulin secretion rate (BasalISR) (141,425] (figures in 2^nd^ column), high levels of visceral adipose tissue (VAT) (8.76,14.5] (figures in 3^rd^ column) and high levels of both (figures in 4^th^ column). Continuous variables were discretized with Hartemink’s method^17^.

### Bayesian network and Mendelian randomization assessment of the Twin-cycle model (IMI DIRECT)

Taylor’s Twin-cycle model, a hypothesized mechanistic model linking NAFLD and T2D, postulates that chronic energy excess leads to accumulation of fat in the liver and subsequently the pancreas, which drives dysglycemia and diabetes. Here, we made a comparison of two models both using the same set of variables: i) an unsupervised BN and ii) the model hypothesized by Taylor. The bidirectional two-sample MR analyses was also used to test causal relationships between all possible combinations of selected variables. The unsupervised BN constructed using the variables of the Twin-cycle model is presented in Fig 3A, where panel B reports the strength and directional probabilities of the network’s arcs. The arcs, mimicking the original Twin-cycle model, include fasting glucose and OGIS towards BasalISR, from glucose sensitivity and OGIS to 2-hour glucose, and from BasalISR to liver fat. Among those arcs not following the Twin-cycle model, TG to liver fat, glucose sensitivity to OGIS and fasting glucose to glucose sensitivity had weak directional probabilities (0.57, 0.66 and 0.60, respectively), which may help explain why the metabolic network predicted in our model is not fully compatible with the network hypothesized in the Twin-cycle model. Through the conducted MR analyses, only the association of fasting glucose to BasalISR could be tested, yet it did not reach the multiple testing corrected significance threshold (P_FDRcorrected_ =0.59).

**Fig 3.**
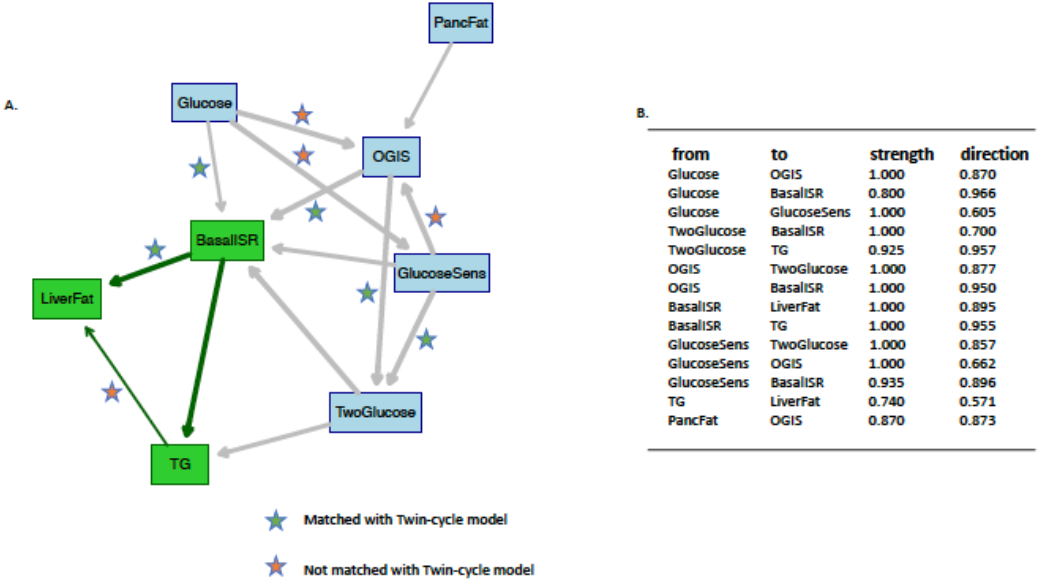
**Panel A:** averaged Bayesian network of the bootstrapped samples among the variables of the Twin-cycle model using data from IMI DIRECT combined cohorts (data are inverse normal transformed, n=1264). **Panel B:** strength and directional probabilities of the arcs presented in the Bayesian network of the panel A. Nodes in green highlight the Markov blanket of liver fat that includes the nodes with adequate information to stand as a separate Bayesian network. BasalISR, insulin secretion at the beginning of the oral glucose tolerance test/ mixed-meal tolerance test; Glucose, fasting glucose from venous plasma samples; GlucoseSens, glucose sensitivity, slope of the dose–response relating insulin secretion to glucose concentration; OGIS, oral glucose insulin sensitivity index according to the method of Mari et al. ^16^; PancFat, pancreas fat; TG, fasting triglycerides; TwoGlucose, 2-hour glucose after oral glucose tolerance test/mixed-meal tolerance test; TwoInsulin, 2-hour insulin.

### Bayesian network and Mendelian randomization in the UK Biobank cohort

As with the IMI DIRECT, we replicated the BN analyses in the UK Biobank dataset where data permitted. We first checked the heatmap cluster among the inverse normal transformed variables (S6 Fig A). The graphical connection of the variables with Pearson correlation ≥0.4 is also presented in S6 Fig B, identifying a cluster of abdominal fat, BMI, waist circumference and liver enzymes. As with the IMI DIRECT analyses, we performed network averaging of the bootstrapped BN samples in the UK Biobank dataset. Table 3 reports the conditional density and parameters for each node of the UK Biobank’s constructed BN. Panel A in Fig 4 shows the averaged BN and panel B illustrates the cumulative distribution function of the arc strengths. Detailed information of those arcs with strength and directional probabilities equal to or greater than the significant threshold (0.5) is reported in S2 Table, along with the available MR results.

**Table 3.**
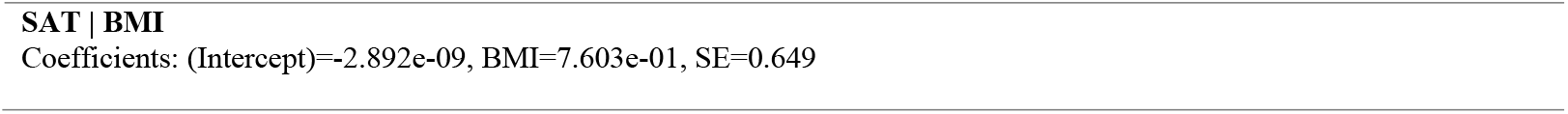
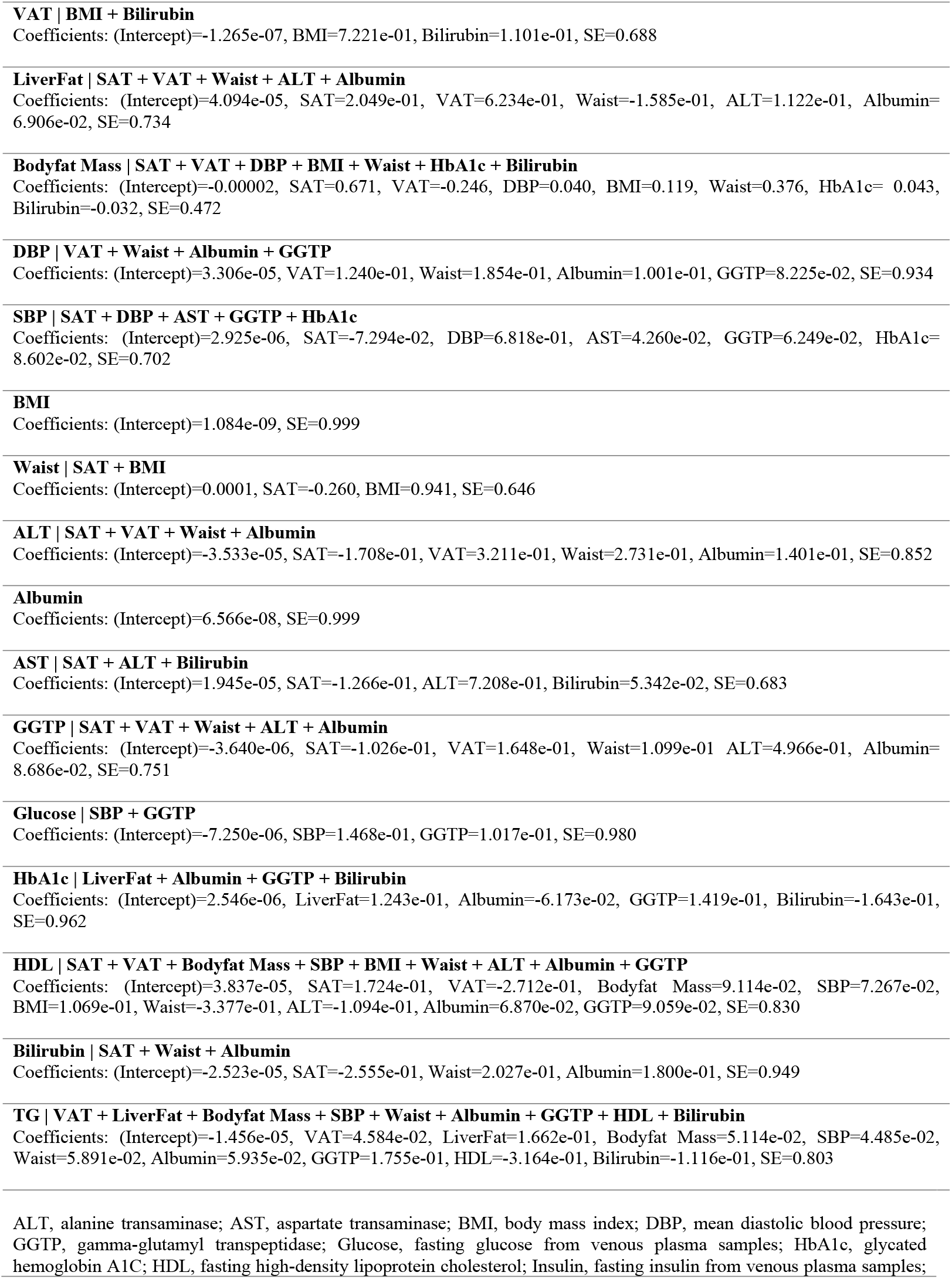
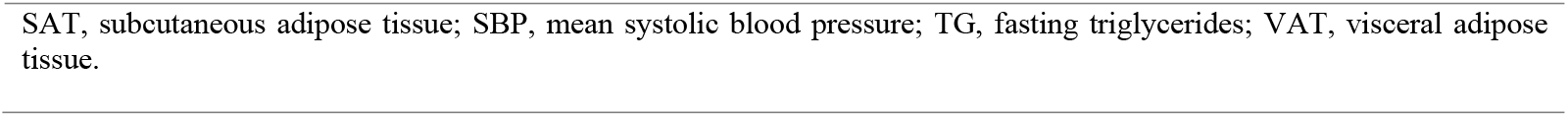
Conditional density and parameters for each node of the constructed average Bayesian network using UK biobank inverse normal transformed data (n=3641). SE: standard error.

**Fig 4.**
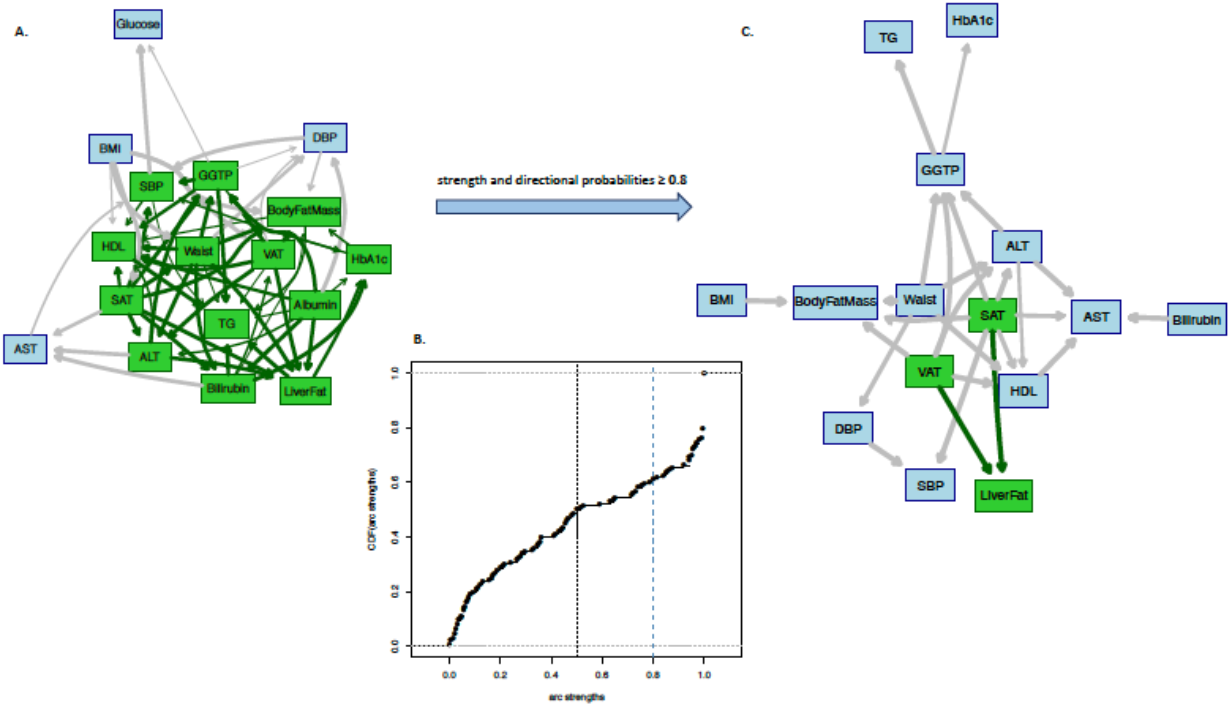
**Panel A:** Averaged Bayesian network of the bootstrapped samples among the variables of UK Biobank (data are inverse normal transformed, n=3641). **Panel B**: Cumulative distribution function of the arc strengths presented in the Bayesian network of Panel A. Black vertical line is the threshold (0.505) learned from data, equal or above which the arcs are identified as statistically significant. **Panel C**: Subset of Bayesian networks in Panel A including only arcs with strength and directional probabilities ≥ 0.8. Nodes in green highlight the Markov blanket of liver fat that includes the nodes with adequate information to stand as a separate Bayesian network. ALT, alanine transaminase; AST, aspartate transaminase; BMI, body mass index; DBP, mean diastolic blood pressure; GGTP, gamma-glutamyl transpeptidase; Glucose, fasting glucose from venous plasma samples; HbA1c, glycated hemoglobin A1C; HDL, fasting high-density lipoprotein cholesterol; Insulin, fasting insulin from venous plasma samples; SAT, subcutaneous adipose tissue; SBP, mean systolic blood pressure; TG, fasting triglycerides; VAT, visceral adipose tissue.

VAT, SAT, waist circumference, ALT and Albumin were identified as the causal parental nodes, and the effect child nodes were HbA1c and TG. The Markov blanket of liver fat included VAT, SAT, waist circumference, whole-body fat mass, systolic blood pressure (SBP), ALT, GGTP, HDL, TG, Albumin, Bilirubin and HbA1c. Fig 4, Panel C shows the BN with only the strong arcs (both strength and directional probabilities ≥ 0.8), where only VAT and SAT remained as the causal nodes of liver fat. We then checked the averaged BN arcs with the two-sample MR results where possible.

Among these arcs, the consistent associations between the MR and BN were waist circumference to BMI (as described above for IMI DIRECT, results shown in SD units, otherwise stated). The association of HbA1c, preceded by fasting glucose (per mmol/L) was also noted as described above. BMI, and waist circumference were causally related to whole-body fat mass (β = 0.76, P_FDRcorrected_= 5.88E-171, P_Egger Intercept_ = 3.4E-01; β = 0.87, P_FDRcorrected_= 4.65E-139, P_Egger Intercept_ = 6.44E-01, respectively). The associations linked with HDL, were those related to adiposity/anthropometry, i.e., waist circumference (as described above), and BMI (β = -0.31, P_FDRcorrected_= 1.79E-10, P_Egger Intercept_ = 2.97E-01; β = -0.25, P_FDRcorrected_= 3.96E-07, P_Egger Intercept_ = 8.84E-01, respectively); moreover, ALT was associated with HDL (β = -0.28, P_FDRcorrected_= 3.61E-03, P_Egger Intercept_ = 4.84E-01). Waist circumference, HDL, GGTP, and whole-body fat mass were all associated with TG (β = 0.22, P_FDRcorrected_= 1.59E-08, P_Egger Intercept_ =2.97E-01; β = -0.34, P_FDRcorrected_= 1.81E-08, P_Egger Intercept_ =1.04E-01; β = 0.21, P_FDRcorrected_= 2.83E-05, P_Egger Intercept_ = 1.02E-01; β = 0.12, P_FDRcorrected_= 1.73E-03, P_Egger Intercept_ = 1.09E-01, respectively).

### Subgroup analysis: diabetes vs. non-diabetes - female vs. male (in UK Biobank)

As with the IMI DIRECT analyses reported above, we built BNs separately in participants diagnosed with T2D (S7 Fig, lower panel) and without diabetes (S7 Fig, upper panel). VAT, SAT and albumin were identified as the parental nodes of liver fat in both groups; ALT and waist circumference were only present as the parents in the group without T2D (S7 Fig panels A and C). The subset of these BNs with arcs having strengths and directional probabilities ≥0.8 left only VAT as the causal node for liver fat in the group with T2D and VAT and SAT in the group without T2D (S7 Fig panels B and D). We then continued with the sensitivity analysis by gender, as illustrated in S8 Fig. In the female’s BN, VAT, TG, ALT, whole-body fat mass, HbA1c and BMI were the causal parental nodes for liver fat, whereas in the male’s BN, VAT, whole-body fat mass and Albumin were the causal variables (S8 Fig, panel A and C). These BNs with extra restrictions on the strength and directional probabilities above or equal to 0.8 resulted in only VAT as the causal parental node for liver fat (S8 Fig, panel B and D). The Pearson correlation and the heatmap cluster analyses, performed in the sensitivity analyses of the UK Biobank resulted in similar clusters to the combined cohorts’ one (see S6, S9 and S10 Figs). Detailed information of arcs with strength and directional probabilities above or equal to the UK Biobank BNs’ thresholds (0.49, 0.5, 0.49 and 0.48 in the non-diabetes, diabetes, female and male BNs, respectively) are reported in S2 Table with their corresponding MR results.

### Bayesian networks and the posterior probabilistic inference (UK Biobank)

Owing to their role as causal parental nodes for liver fat in the UK Biobank BNs, high levels of VAT and SAT (see S3 Table) were investigated for their conditional effects on posterior probabilities of liver fat levels. Fig 5 summarizes unconditional and posterior conditional probabilities of liver fat derived using complete case data, diabetes, non-diabetes, female and male subgroups of the UK Biobank BNs. A statistically significant increase was observed after conditioning on high levels of VAT and SAT in the unconditional probability of high liver fat level on the random observations generated by the BNs from the complete case data, 37% increase after conditioning on VAT, 18% increase after conditioning on SAT, and 41% after conditioning on both. Similarly, diabetes, non-diabetes, female and male groups had substantial increments on the high liver fat level probability after conditioning on high levels of VAT, SAT and the combined effect. The probability of having high liver fat after conditioning on high levels of VAT and SAT was 0.70, 0.97, 0.79, 0.74 and 0.72 in the complete case data, diabetes, non-diabetes, female and male groups of the UK Biobank constructed BNs.

**Fig 5.**
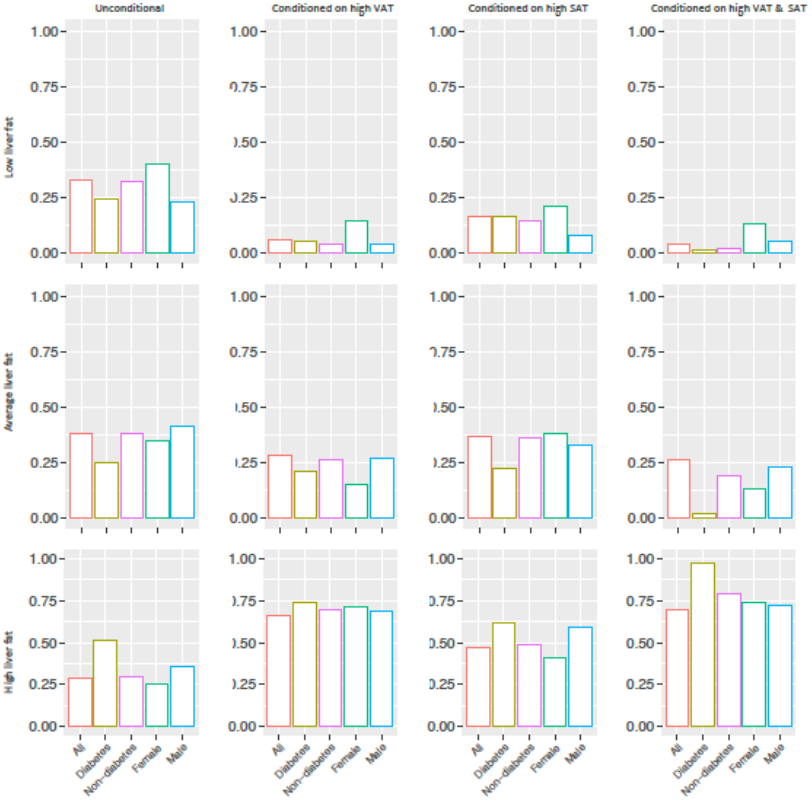
Unconditional and posterior conditional probabilities of liver fat on the combined (n=3641), diabetes (n=104), non-diabetes (n=3531), female (n=1921) and male (n=1720) groups of the UK biobank Bayesian networks (data are inverse normal transformed). Y axis represents the unconditional and posterior conditional probabilities of low (0,1.438], average (1.438,3.619] or high (3.619,46.049] levels of liver fat in rows. The posterior probabilities were obtained after conditioning on high levels of visceral adipose tissue (VAT) (5.958,14.408] (figures in 2^nd^ column), high levels of subcutaneous adipose tissue (SAT) (7.808,23.476] (figures in 3^rd^ column) and high levels of both (figures in 4^th^ column). Continuous variables were discretized with Hartemink’s method^17^.

## DISCUSSION

We interrogated the putative causal pathways between metabolic processes and liver fat accumulation through Bayesian models and a series of bidirectional MR analyses. BNs fitted to each dataset separately suggested that VAT and BasalISR (only available in IMI DIRECT) causally affect liver fat accumulation. Reassuringly, VAT and BasalISR were also identified as the strongest causal determinants through different subgroup analyses as the closest determinants of liver fat accumulation. In addition, the probabilistic inference analyses showed a substantial increase in the posterior probability of having high levels of liver fat after conditioning on VAT and BasalISR (∼30%-40%). Furthermore, the MR analysis suggests that several nominal directional associations between hepatic biomarkers, glycemic and adiposity measures are likely to be causal (summarized in S1 Table).

The epidemiological association of NAFLD and T2D has been widely reported, yet it is unclear if this relationship is causal, and, if so, whether it is uni- or bi-directional, and what the underlying mechanisms might be. MR has been widely applied to epidemiological datasets to help minimize confounding and reverse-causation that are common weaknesses of such studies^18,19^. In a recent study from Liu et.al, the causal relationships between NAFLD, T2D and obesity were explored using a bidirectional 2-sample MR analysis in UK Biobank^9^. Their results showed that genetically-determined NAFLD causes T2D and central obesity, whereas genetically-determined T2D, overall and central obesity cause metabolic NAFLD. Another smaller-scale MR analysis^20^ suggested the causal role of hepatic fat accumulation in the development of chronic liver disease and insulin resistance. The hepatic enzymes ALT and AST were found to be causally associated with T2D through bidirectional MR analyses that assessed the causal relationship of NAFLD and T2D^21^, something that was not observed in our analyses. BN is an established causal inference method^22,23^, yet it has rarely been utilized to study metabolic traits^24^. Within these rare exceptions, a reciprocal association between NAFLD and metabolic syndrome was suggested through a simplified BN applied to a Chinese cohort^25^. Aiming to predict complications of T2D, BN models were built from physiological risk factors of the disease^26^. BN have also been used to study relationships between risk factors for T2D in European-ancestry cohorts^27^.

Through the main BN analyses reported here, BasalISR was the strongest causal driver of liver fat accumulation. This may be explained by hepatic insulin sensitivity (not measured in IMI DIRECT nor UK Biobank) or as a direct effect of BasalISR as shown in a recent study by van Vliet et al. ^28^ where a clinical association between obesity and basal insulin secretion rate was observed even in the absence of insulin resistance^28^. Our analyses also showed that in people with diabetes, although BasalISR was an important factor, VAT had a bigger causal impact on liver fat accumulation (Fig 2- compare 2^nd^ and 3^rd^ figures in the last row). In the UK Biobank analyses, where BasalISR was unavailable, VAT was the strongest determinant of fatty liver. This, combined with findings in the diabetes sub-cohort of IMI DIRECT, suggests that VAT is likely to be a strong causal determinant of fatty liver, especially when diabetes has progressed, and insulin secretion is diminished. Our findings were also consistent with other studies that show that excess visceral adiposity is an important predictor of NAFLD, much more so than subcutaneous adiposity^29^ (Fig 5- compare 2^nd^ and 3^rd^ figures in the last row).

In addition to using BNs for causal inference, we used MR to validate the factors involved in the metabolic processes and liver fat pathways. We found that glycemic traits were at least nominally associated with liver enzymes (i.e., AST and GGTP), but the reciprocal effect was not detected; however, AST was causally related with insulin sensitivity, though this was not detected in pleiotropy/outlier sensitivity analyses. Although there are no robust genetic instruments for VAT and SAT^30^, which would be necessary for an MR analysis assessing these traits, this could be done for whole-body fat mass. Here, the MR analysis supports a causal association of whole-body fat mass with insulin sensitivity, consistent with the findings from the BNs. Moreover, larger waist circumference and higher BMI were causally associated with fasting insulin levels, yet there was no evidence of an effect of these traits on insulin secretion.

In a recent analysis from IMI DIRECT consortium^31^, we used structural equation modelling to test the hypothesis that physical activity affects glucose regulation in a manner consistent with the Twin-cycle hypothesis^6,7^. Here, we constructed a BN that was agnostic to the hypothesized Twin-cycle model and showed good, albeit not complete, agreement between the two models, providing further validation of the Twin-cycle hypothesis. Note that BN is acyclical, thus preventing complete recapitulation of the Twin cycle hypothesis using this approach. Moreover, instead of hepatic insulin sensitivity (as used in the Twin-cycle model), OGIS, the insulin sensitivity index, was used as a proxy, which might explain some of the observed differences. Furthermore, there are currently no genetic instruments available that adequately characterize pancreatic fat^32^, thus preventing its inclusion in our MR analyses.

Although BN and MR have key limitations, they are largely orthogonal methods and their combination provides a fairly robust causal inference framework. To avoid any overlap in the two samples utilized in MR analysis (exposure and outcome), we restricted the analyses primarily to the non-UK Biobank repositories. However, when we had both exposure and outcome derived from the same repositories, we considered UK Biobank for one of them. Low statistical power can also be a problem in MR analyses, owing to the conditional nature of the calculations performed. Specifically, MR depends on the proportion of variance in a phenotypic trait that is explained by the instrumental variables (SNPs) used in the GWAS; thus, a limited number of instrumental variables is unlikely to capture a true effect for complex exposures. Moreover, established methodological caveats of MR, such as pleiotropy and heterogeneity, were assessed to effectively disentangle the effect of each trait; however, we cannot completely rule out that our instrumental variables have other phenotypic effects. We adopted significance criteria with a conservative p-value threshold; the concordance with two methods, and no statistical evidence of pleiotropy. Within the same context, utilizing BNs for causal inference needs careful consideration, the assumption being there are no latent variables in the model. Here, we focused on probabilistic inference and computed the posterior probability of liver fat from the observed BNs after conditioning on different pieces of evidence (high levels of BasalISR, VAT and/or SAT). It is noteworthy that the constructed BNs should be considered as probable causal models, since they are derived from available data and knowledge.

Another limitation of our approach was that to learn the structure and parameters of the BNs, a complete case analysis was required, which diminished the sample sizes both in UK Biobank and IMI DIRECT considerably. Amongst those from UK Biobank, liver iron and liver inflammation factors were two interesting variables that we elected not to include in the BN analyses, due to high missing rates (∼70%). However, as sensitivity analyses, we tried building BN in the subset including liver iron and liver inflammation factor, and it had a downstream association from liver fat, similar to the IMI DIRECT network. Subsetting the UK Biobank datasets to only those diagnosed with T2D left very few individuals for the complete case analyses. As such, the BN model for the diabetic subset is sparse and possibly underpowered. In a recent study, Scutari reviewed how BNs can model data with incomplete observations^33^, which can be a focus for future research. Multicollinearity, where input variables are highly correlated, can be an issue in some pathway analyses. However, the type of structural learning in a BN makes it robust against such limitation. This property makes BN an intuitive visual tool to express associations among several variables in a complex network. We did not apply any force node in building the BNs and they were built unsupervised. However, semi-supervised networks feeding with known directional associations (e.g., from previous MR analyses) can be considered as future work. Expansion of the Bayesian workspace to omics^34^ would be a logical extension of the current work, especially considering the multi-omics data (genetic, transcriptomic, proteomic and metabolomic) available in IMI DIRECT consortium.

In summary, the combination of BN and MR analyses deployed here provided a powerful causal inference framework through which a causal model of metabolic homeostasis in fatty liver disease could be developed and validated. The key findings are that insulin secretion rate and visceral adiposity are major causal drivers of fatty liver, until, at least, diabetes develops, at which point visceral adiposity becomes the dominant causal agent. These findings may aid the targeted prevention of fatty liver disease.

## MATERIALS AND METHODS

### IMI DIRECT cohorts and measures

We utilized data from the IMI DIRECT consortium, prospective cohort studies of 795 adults with T2D and 2234 without the disease. Written informed consent was provided by all participants at enrollment and the regional research ethics committees of each clinical study center have approved the study protocol separately^35,36^. The participants were extensively assessed with measures focused on glycemia, insulin dynamics, organ-specific adiposity, serological biomarkers, lifestyle, anthropometry and other clinical features. Frequently sampled mixed-meal tolerance tests (MMTTs) and 75 g frequently sampled oral glucose tolerance tests (fsOGTTs) were carried out in the diabetes and non-diabetes cohorts, respectively. Basal insulin secretion rate (BasalISR) was estimated as the product of insulin clearance and fasting plasma insulin as defined elsewhere^37^. Adiposity, including liver fat, pancreas fat, visceral fat and subcutaneous fat, was assessed using a T2*-based multiecho technique from MRI scans^38^. An overview of participant characteristics is shown in S3 Table in the non-diabetes, diabetes and combined cohorts of the IMI DIRECT. More details of the study design and the core characteristics are provided elsewhere^35,36^.

### UK Biobank cohort and measures

UK Biobank is a prospective cohort study with more than 500,000 adults aged 37–73 years and recruited between 2006 and 2010^39^. The current analysis was conducted using data obtained via the UK Biobank Access Application process (project number 18274). We used this dataset for both modelling and replication of the primary findings reported herein form the IMI DIRECT cohorts. UK Biobank and IMI DIRECT have the same protocol and procedure for quantification of the MRI-derived abdominal fat^40^. The field numbers for the UK Biobank variables and an overview of UK Biobank participants’ characteristics are shown in S3 Table.

### Bayesian Network analysis

We utilized BNs to build the graphical models for inferring causal pathways. BNs are probabilistic graphical models that are built from joint probability distributions of random variables using Bayesian inference. The network *structure* is depicted graphically by directed acyclic graphs (DAGs), where nodes represent the random variables with directed arcs affirming the conditional dependencies and missing arcs defining the conditional independencies in the model^41-43^.

Building a BN can be summarized into a two-step learning process (equation 1); structural learning where the network structure is learned from data and parameter learning where local distributions and parameters are inferred from the learned structure in the first step:

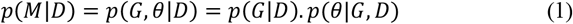

Where *p*(*M*|*D*) is the likelihood of the BN model according to the structure G and parameter *θ* given data D. The structural learning in our analyses has been undertaken by the *score-based* approach, as it deals better with the small sample size and the possible noise in the data^44^. The heuristic search using the hill-climbing technique^45^ was applied, searching towards the structure with the maximum goodness of fit score. Scoring of the BN structures was done using the *Bayesian Information Criterion* (BIC) score method as follows:

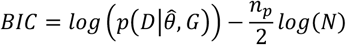

where, *p*(*D*|*θ,G*) is the probability of data given parameters and the graph, *n*_*p*_ is the number of parameters with *N* denoting the sample size. We elected the BIC method as it penalizes networks with many arcs, leading to a simpler graph with fewer false positives^45^.

Numeric variables were all inverse normalized to fit a Gaussian BN with a multivariate normal distribution^46,47^. Such networks require that the nodes are normally distributed to function properly. Furthermore, to attain a stable structure from data, which is resistance to the network perturbation, we performed model averaging. As such, data were resampled using bootstrapping, following which a separate structure was built for each bootstrap sample, and the averaged network was then constructed based on the frequency of the possible arcs amongst all networks.

The *Markov condition* is an assumption that must be satisfied when making the BNs, which states a node can be conditionally independent of the whole network given its Markov blanket (the set of nodes that includes all the information required for separating it from the rest of graph)^48^. In our networks, the Markov blanket of the liver fat node, which includes the upstream (parent) nodes, the downstream (children) nodes and other parents of those children, were derived. After learning the structure of the network, the parameters were fitted using their maximum likelihood estimate.

To further investigate a constructed BN after its structure (*G*) and parameter (*θ*) learning, *queries* on the variable of interest (*X*), known as *event*, can be defined. Here, we focused on liver fat as our event and computed its posterior probability after conditioning on an *evidence* (*E*) to further understand its causal effect on liver fat, *p*(*X*|*E,G, θ*). To compute the conditional posterior probabilities, variables were required to be discrete. To discretize the continues variables, Hartemink’s method was deployed which helps to preserve the initial dependencies among the variables while defining the intervals^17^.

### Mendelian randomization analysis

The MR approach has gained popularity because it is less susceptible to confounding and reverse causation than most other observational studies^49,50^, and is often likened to a randomized controlled trial (RCT) owing to the random allocation of most confounding variables across the levels of the exposure instrument (genotype). Specifically, the random assortment of alleles during meiosis allows exploration of causal relationships between various traits and diseases under the assumption that genetic variants [known as instrumental variables (IV)] are suitable proxies for given risk factors, moreover, the instruments should influence the outcome only through the exposure and not through any other confounding^18^. Bidirectional MR involves assessment of the reciprocal exposure-outcome relationship to assess the directionality in causation. Thus, a systematic bidirectional MR study leveraging the latest public GWAS data would help triangulate causality of directed arcs in BN analysis.

Two-sample MR analysis was deployed, where each trait was investigated for potential causal associations. Between the selected metabolic traits, we assessed bidirectionally, i.e. used as exposure and as outcome^50^, we leveraged from the latest summary statistics from publicly available sources i.e. GWAS catalog^51^ and MR-CIEU^52^. Operationally, not all traits defined in IMI DIRECT or UK Biobank have a corresponding GWAS published. We prioritized GWAS restricted to European population (see S1 Table) based on i) latest release, ii) sample size, and iii) if no published GWAS was obtained or not enough number of independent variants were identified we used the UK Biobank data^53^ from Neale Lab (http://www.nealelab.is/blog/2017/9/11/details-and-considerations-of-the-uk-biobank-gwas). Additionally, in case of sample overlap between two datasets, we preemptively selected one trait from UK Biobank and overlap was calculated with the maximum sample-overlapping rate ^55^, as reported by Liu and colleagues^9^. Instrumental variables were prioritized at GWAS-significance threshold (p-value<5 × 10^−8^) and proxies were used if the genetic variants were in linkage disequilibrium (LD) at *r*^2^ ≥ 0.8 in any of the two-samples, yet, to avoid LD within the instruments, we performed LD-clumping restricted to *r*^2^ < 0.2, a 1000 kb window for the final sets.

We used the inverse variance weighted (IVW) method as the main analysis to estimate the effects of the instrumental variables, when there were sufficient number of genetic variants. Furthermore, we used MR-Egger method to minimize false positive associations. To quantify heterogeneity, horizontal pleiotropy, and detect outliers, we used the MR-Egger intercept, the Q statistic and the MR Pleiotropy Residual Sum and Outlier (MR-PRESSO) global test at p level of >0.05 when appropriate^55^. MR-Egger, as part of the regression, provides a consistent estimate of the causal effect. Moreover, when the InSIDE (Instrument Strength Independent of Direct Effect) assumption holds, the intercept provides an unbiased average pleiotropic effect that should not significantly differ from the null, yet, when this is unmet, its value represents an estimate of the directional horizontal pleiotropy^56^. We considered MR findings to be statistically significant if: i) the causal association amongst IVW, and Egger were directionally concordant, ii) the IVW (main method) passed the FDR corrected^13^ threshold (p<0.05) for multiple-testing, and iii) no statistical evidence of heterogeneity and/or pleiotropy (PEgger Intercept, and PMR-PRESSOGlobal p>0.05).

All the statistical analyses were undertaken using R software version 3.6.2^57^, the BNs were built using the *bnlearn* package^58,59^ and the MR analysis using the *TwoSampleMR*^52^ and MR-PRESSO packages^55^. Variables from IMI DIRECT were adjusted for center effect in a linear model including each variable per model. The residuals were then extracted from these models and were ranked normalized to fulfill the Gaussian BN assumption. The distribution of the IMI DIRECT and UK Biobank variables prior to transformation is depicted in S11 and S12 Figs, respectively.

## Data Availability

Authors agree to make data and materials supporting the results or analyses presented in their paper available upon reasonable request

## SUPPLEMENTARY INFORMATION

**S1 Fig**. A. Heatmap cluster among the variables of IMI DIRECT combined cohorts (data are inverse normal transformed, n=1070). B. The graphical connection of the variables with Pearson correlation equal or above 0.4.

OGIS, oral glucose insulin sensitivity index according to the method of Mari et al. ^16^; Clins, mean insulin clearance during the oral glucose tolerance test/mixed meal tolerance test, calculated as (mean insulin secretion)/(mean insulin concentration); HDL, fasting high-density lipoprotein cholesterol; GlucoseSens, glucose sensitivity, slope of the dose–response relating insulin secretion to glucose concentration; PancIron, pancreas iron; PancFat, pancreas fat; SBP, mean systolic blood pressure; DBP, mean diastolic blood pressure; Glucagonmin0, fasting glucagon concentration; TotGLP1min0, concentration of fasting total GLP-1 in plasma; HbA1c, glycated hemoglobin A1C; Glucose, fasting glucose from venous plasma samples; TwoGlucose, 2-hour glucose after oral glucose tolerance test/mixed-meal tolerance test; GGTP, gamma-glutamyl transpeptidase; AST, aspartate transaminase; ALT, alanine transaminase; SAT, subcutaneous adipose tissue; BMI, body mass index; TG, fasting triglycerides; VAT, visceral adipose tissue; TwoInsulin, 2-hour insulin; Insulin, fasting insulin from venous plasma samples; BasalISR, insulin secretion at the beginning of the oral glucose tolerance test/ mixed-meal tolerance test.

**S2 Fig**. Averaged Bayesian network of the bootstrapped samples among the variables of IMI DIRECT cohorts, panel A-B for non-diabetic (n=787) and panel C-D for diabetic (n=283) cohorts. Data are inverse normal transformed and the Bayesian networks in panels B and D show only the arcs with strength and directional probabilities ≥ 0.8.

OGIS, oral glucose insulin sensitivity index according to the method of Mari et al.^16^; Clins, mean insulin clearance during the oral glucose tolerance test/mixed meal tolerance test, calculated as (mean insulin secretion)/(mean insulin concentration); HDL, fasting high-density lipoprotein cholesterol; GlucoseSens, glucose sensitivity, slope of the dose–response relating insulin secretion to glucose concentration; PancIron, pancreas iron; PancFat, pancreas fat; SBP, mean systolic blood pressure; DBP, mean diastolic blood pressure; Glucagonmin0, fasting glucagon concentration; TotGLP1min0, concentration of fasting total GLP-1 in plasma; HbA1c, glycated hemoglobin A1C; Glucose, fasting glucose from venous plasma samples; TwoGlucose, 2-hour glucose after oral glucose tolerance test/mixed-meal tolerance test; GGTP, gamma-glutamyl transpeptidase; AST, aspartate transaminase; ALT, alanine transaminase; SAT, subcutaneous adipose tissue; BMI, body mass index; TG, fasting triglycerides; VAT, visceral adipose tissue; TwoInsulin, 2-hour insulin; Insulin, fasting insulin from venous plasma samples; BasalISR, insulin secretion at the beginning of the oral glucose tolerance test/ mixed-meal tolerance test.

**S3 Fig**. Averaged Bayesian network of the bootstrapped samples among the variables of IMI DIRECT cohorts, panel A-B for female (n=293) and panel C-D for male (n=831) groups. Data are inverse normal transformed and the Bayesian networks in panels B and D show only the arcs with strength and directional probabilities ≥ 0.8.

OGIS, oral glucose insulin sensitivity index according to the method of Mari et al.^16^; Clins, mean insulin clearance during the oral glucose tolerance test/mixed meal tolerance test, calculated as (mean insulin secretion)/(mean insulin concentration); HDL, fasting high-density lipoprotein cholesterol; GlucoseSens, glucose sensitivity, slope of the dose–response relating insulin secretion to glucose concentration; PancIron, pancreas iron; PancFat, pancreas fat; SBP, mean systolic blood pressure; DBP, mean diastolic blood pressure; Glucagonmin0, fasting glucagon concentration; TotGLP1min0, concentration of fasting total GLP-1 in plasma; HbA1c, glycated hemoglobin A1C; Glucose, fasting glucose from venous plasma samples; TwoGlucose, 2-hour glucose after oral glucose tolerance test/mixed-meal tolerance test; GGTP, gamma-glutamyl transpeptidase; AST, aspartate transaminase; ALT, alanine transaminase; SAT, subcutaneous adipose tissue; BMI, body mass index; TG, fasting triglycerides; VAT, visceral adipose tissue; TwoInsulin, 2-hour insulin; Insulin, fasting insulin from venous plasma samples; BasalISR, insulin secretion at the beginning of the oral glucose tolerance test/ mixed-meal tolerance test.

**S4 Fig**. Heatmap cluster and the graphical connection of the IMI DIRECT variables with Pearson correlation equal or above 0.4, panel A-B for non-diabetic (n=787) and panel C-D for diabetic (n=283) cohorts. Data are inverse normal transformed.

OGIS, oral glucose insulin sensitivity index according to the method of Mari et al^16^; Clins, mean insulin clearance during the oral glucose tolerance test/mixed meal tolerance test, calculated as (mean insulin secretion)/(mean insulin concentration); HDL, fasting high-density lipoprotein cholesterol; GlucoseSens, glucose sensitivity, slope of the dose–response relating insulin secretion to glucose concentration; PancIron, pancreas iron; PancFat, pancreas fat; SBP, mean systolic blood pressure; DBP, mean diastolic blood pressure; Glucagonmin0, fasting glucagon concentration; TotGLP1min0, concentration of fasting total GLP-1 in plasma; HbA1c, glycated hemoglobin A1C; Glucose, fasting glucose from venous plasma samples; TwoGlucose, 2-hour glucose after oral glucose tolerance test/mixed-meal tolerance test; GGTP, gamma-glutamyl transpeptidase; AST, aspartate transaminase; ALT, alanine transaminase; SAT, subcutaneous adipose tissue; BMI, body mass index; TG, fasting triglycerides; VAT, visceral adipose tissue; TwoInsulin, 2-hour insulin; Insulin, fasting insulin from venous plasma samples; BasalISR, insulin secretion at the beginning of the oral glucose tolerance test/ mixed-meal tolerance test.

**S5 Fig**. Heatmap cluster and the graphical connection of the IMI DIRECT variables with Pearson correlation equal or above 0.4, panel A-B for female (n=293) and panel C-D for male (n=831) groups. Data are inverse normal transformed.

OGIS, oral glucose insulin sensitivity index according to the method of Mari et al.^16^; Clins, mean insulin clearance during the oral glucose tolerance test/mixed meal tolerance test, calculated as (mean insulin secretion)/(mean insulin concentration); HDL, fasting high-density lipoprotein cholesterol; GlucoseSens, glucose sensitivity, slope of the dose–response relating insulin secretion to glucose concentration; PancIron, pancreas iron; PancFat, pancreas fat; SBP, mean systolic blood pressure; DBP, mean diastolic blood pressure; Glucagonmin0, fasting glucagon concentration; TotGLP1min0, concentration of fasting total GLP-1 in plasma; HbA1c, glycated hemoglobin A1C; Glucose, fasting glucose from venous plasma samples; TwoGlucose, 2-hour glucose after oral glucose tolerance test/mixed-meal tolerance test; GGTP, gamma-glutamyl transpeptidase; AST, aspartate transaminase; ALT, alanine transaminase; SAT, subcutaneous adipose tissue; BMI, body mass index; TG, fasting triglycerides; VAT, visceral adipose tissue; TwoInsulin, 2-hour insulin; Insulin, fasting insulin from venous plasma samples; BasalISR, insulin secretion at the beginning of the oral glucose tolerance test/ mixed-meal tolerance test.

**S6 Fig**. A. Heatmap cluster among the variables of the UK Biobank (data are inverse normal transformed, n=3641). B. The graphical connection of the variables with Pearson correlation equal or above 0.4.

ALT, alanine transaminase; AST, aspartate transaminase; BMI, body mass index; DBP, mean diastolic blood pressure; GGTP, gamma-glutamyl transpeptidase; Glucose, fasting glucose from venous plasma samples; HbA1c, glycated hemoglobin A1C; HDL, fasting high-density lipoprotein cholesterol; Insulin, fasting insulin from venous plasma samples; SAT, subcutaneous adipose tissue; SBP, mean systolic blood pressure; TG, fasting triglycerides; VAT, visceral adipose tissue.

**S7 Fig**. Averaged Bayesian network of the bootstrapped samples among the variables of the UK Biobank, panel A-B for non-diabetic (n=3531) and panel C-D for diabetic (n=104) cohorts. Data are inverse normal transformed and the Bayesian networks in panels B and D show only the arcs with strength and directional probabilities ≥ 0.8.

ALT, alanine transaminase; AST, aspartate transaminase; BMI, body mass index; DBP, mean diastolic blood pressure; GGTP, gamma-glutamyl transpeptidase; Glucose, fasting glucose from venous plasma samples; HbA1c, glycated hemoglobin A1C; HDL, fasting high-density lipoprotein cholesterol; Insulin, fasting insulin from venous plasma samples; SAT, subcutaneous adipose tissue; SBP, mean systolic blood pressure; TG, fasting triglycerides; VAT, visceral adipose tissue.

**S8 Fig**. Averaged Bayesian network of the bootstrapped samples among the variables of the UK Biobank, panel A-B for female (n=1921) and panel C-D for male (n=1720) groups. Data are inverse normal transformed and the Bayesian networks in panels B and D show only the arcs with strength and directional probabilities ≥ 0.8.

ALT, alanine transaminase; AST, aspartate transaminase; BMI, body mass index; DBP, mean diastolic blood pressure; GGTP, gamma-glutamyl transpeptidase; Glucose, fasting glucose from venous plasma samples; HbA1c, glycated hemoglobin A1C; HDL, fasting high-density lipoprotein cholesterol; Insulin, fasting insulin from venous plasma samples; SAT, subcutaneous adipose tissue; SBP, mean systolic blood pressure; TG, fasting triglycerides; VAT, visceral adipose tissue.

**S9 Fig**. Heatmap cluster and the graphical connection of the UK Biobank variables with Pearson correlation equal or above 0.4, panel A-B for non-diabetic (n=3531) and panel C-D for diabetic (n=104) groups. Data are inverse normal transformed.

ALT, alanine transaminase; AST, aspartate transaminase; BMI, body mass index; DBP, mean diastolic blood pressure; GGTP, gamma-glutamyl transpeptidase; Glucose, fasting glucose from venous plasma samples; HbA1c, glycated hemoglobin A1C; HDL, fasting high-density lipoprotein cholesterol; Insulin, fasting insulin from venous plasma samples; SAT, subcutaneous adipose tissue; SBP, mean systolic blood pressure; TG, fasting triglycerides; VAT, visceral adipose tissue.

**S10 Fig**. Heatmap cluster and the graphical connection of the UK Biobank variables with Pearson correlation equal or above 0.4, panel A-B for female (n=1921) and panel C-D for male (n=1720) groups. Data are inverse normal transformed.

ALT, alanine transaminase; AST, aspartate transaminase; BMI, body mass index; DBP, mean diastolic blood pressure; GGTP, gamma-glutamyl transpeptidase; Glucose, fasting glucose from venous plasma samples; HbA1c, glycated hemoglobin A1C; HDL, fasting high-density lipoprotein cholesterol; Insulin, fasting insulin from venous plasma samples; SAT, subcutaneous adipose tissue; SBP, mean systolic blood pressure; TG, fasting triglycerides; VAT, visceral adipose tissue.

**S11 Fig**. Distribution of the variables prior to transformation in the IMI DIRECT combined cohorts (n=1070). The blue curve shows the normally transformed distribution of the variables. OGIS, oral glucose insulin sensitivity index according to the method of Mari et al.^16^; Clins, mean insulin clearance during the oral glucose tolerance test/mixed meal tolerance test, calculated as (mean insulin secretion)/(mean insulin concentration); HDL, fasting high-density lipoprotein cholesterol; GlucoseSens, glucose sensitivity, slope of the dose–response relating insulin secretion to glucose concentration; PancIron, pancreas iron; PancFat, pancreas fat; SBP, mean systolic blood pressure; DBP, mean diastolic blood pressure; Glucagonmin0, fasting glucagon concentration; TotGLP1min0, concentration of fasting total GLP-1 in plasma; HbA1c, glycated hemoglobin A1C; Glucose, fasting glucose from venous plasma samples; TwoGlucose, 2-hour glucose after oral glucose tolerance test/mixed-meal tolerance test; GGTP, gamma-glutamyl transpeptidase; AST, aspartate transaminase; ALT, alanine transaminase; SAT, subcutaneous adipose tissue; BMI, body mass index; TG, fasting triglycerides; VAT, visceral adipose tissue; TwoInsulin, 2-hour insulin; Insulin, fasting insulin from venous plasma samples; BasalISR, insulin secretion at the beginning of the oral glucose tolerance test/ mixed-meal tolerance test.

**S12 Fig**. Distribution of the variables prior to transformation in the UK Biobank cohort (n=3641). The blue curve shows the normally transformed distribution of the variables.

ALT, alanine transaminase; AST, aspartate transaminase; BMI, body mass index; DBP, mean diastolic blood pressure; GGTP, gamma-glutamyl transpeptidase; Glucose, fasting glucose from venous plasma samples; HbA1c, glycated hemoglobin A1C; HDL, fasting high-density lipoprotein cholesterol; Insulin, fasting insulin from venous plasma samples; SAT, subcutaneous adipose tissue; SBP, mean systolic blood pressure; TG, fasting triglycerides; VAT, visceral adipose tissue.

**S1 Table**. The 2-sample Mendelian randomization exposure-outcome associations reported per outcome trait.

ALT, alanine transaminase; AST, aspartate transaminase; BMI, body mass index; DBP, mean diastolic blood pressure; GGTP, gamma-glutamyl transpeptidase; HbA1c, glycated hemoglobin A1C; HDL, fasting high-density lipoprotein cholesterol; IDI, insulin disposition index; ISI, insulin sensitivity index; SAT, subcutaneous adipose tissue; SBP, mean systolic blood pressure; TG, fasting triglycerides; VAT, visceral adipose tissue.

**S2 Table**. Information on the strength and directional probabilities of the arcs for the combined, diabetes, non-diabetes, female and male groups of the IMI DIRECT and UK Biobank Bayesian networks. The arcs are limited for those with strength and directional probabilities equal to or greater than the model’s significant threshold. The corresponding Mendelian randomization is reported when available.

**S3 Table**. Characteristics of the IMI DIRECT and UK Biobank in the non-diabetes, diabetes and combined cohorts and the list of the variables used in the analyses with their meanings. All the continuous variables were discretized into three levels (Low, Average and High) with Hartemink’s method for the probabilistic inference analyses and are presented in this table.

## Ethics statements

Approval for the study protocol was obtained from each of the regional research ethics review boards separately (Lund, Sweden: 20130312105459927, Copenhagen, Denmark: H-1-2012-166 and H-1-2012-100, Amsterdam, Netherlands: NL40099.029.12, Newcastle, Dundee and Exeter, UK: 12/NE/0132), and all participants provided written informed consent at enrolment. The research conformed to the ethical principles for medical research involving human participants outlined in the Declaration of Helsinki.

## Acknowledgments

The work leading to this publication has received support from the Innovative Medicines Initiative Joint. Undertaking under grant agreement n°115317 (DIRECT, https://directdiabetes.org/), resources of which are composed of financial contribution from the European Union’s Seventh Framework Programme (FP7/2007-2013) and EFPIA companies’ in kind contribution. We thank all the participants and study center staff in IMI DIRECT for their contribution to the study. We thank all the participants in the UK Biobank. This research was conducted using the UK Biobank resource (application ID: 18274).

