## Supplementary figures and images for "Inferring causal pathways between metabolic processes and liver fat accumulation: an IMI DIRECT study"

### Supplemental figure 1

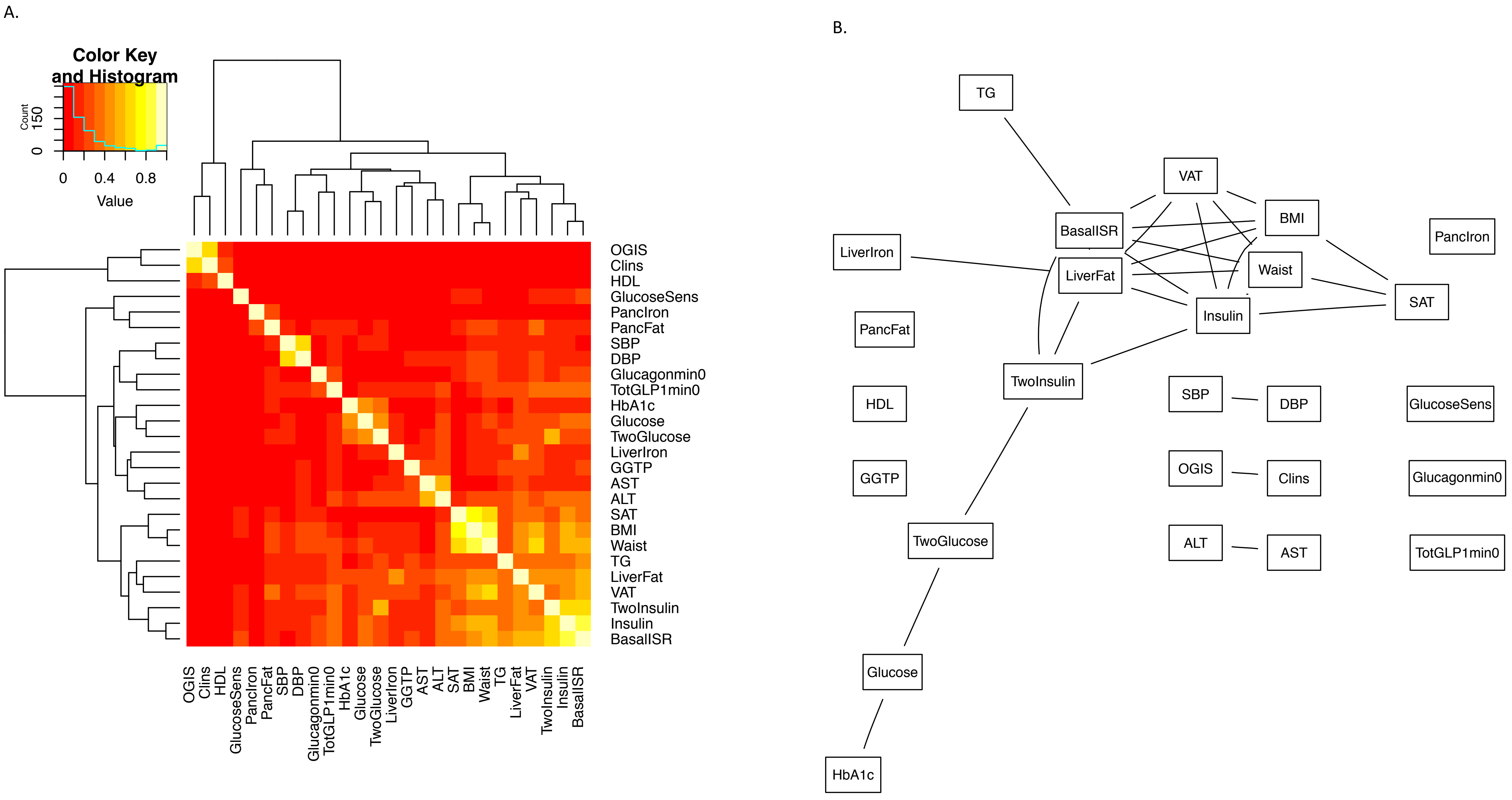

### Supplemental figure 2

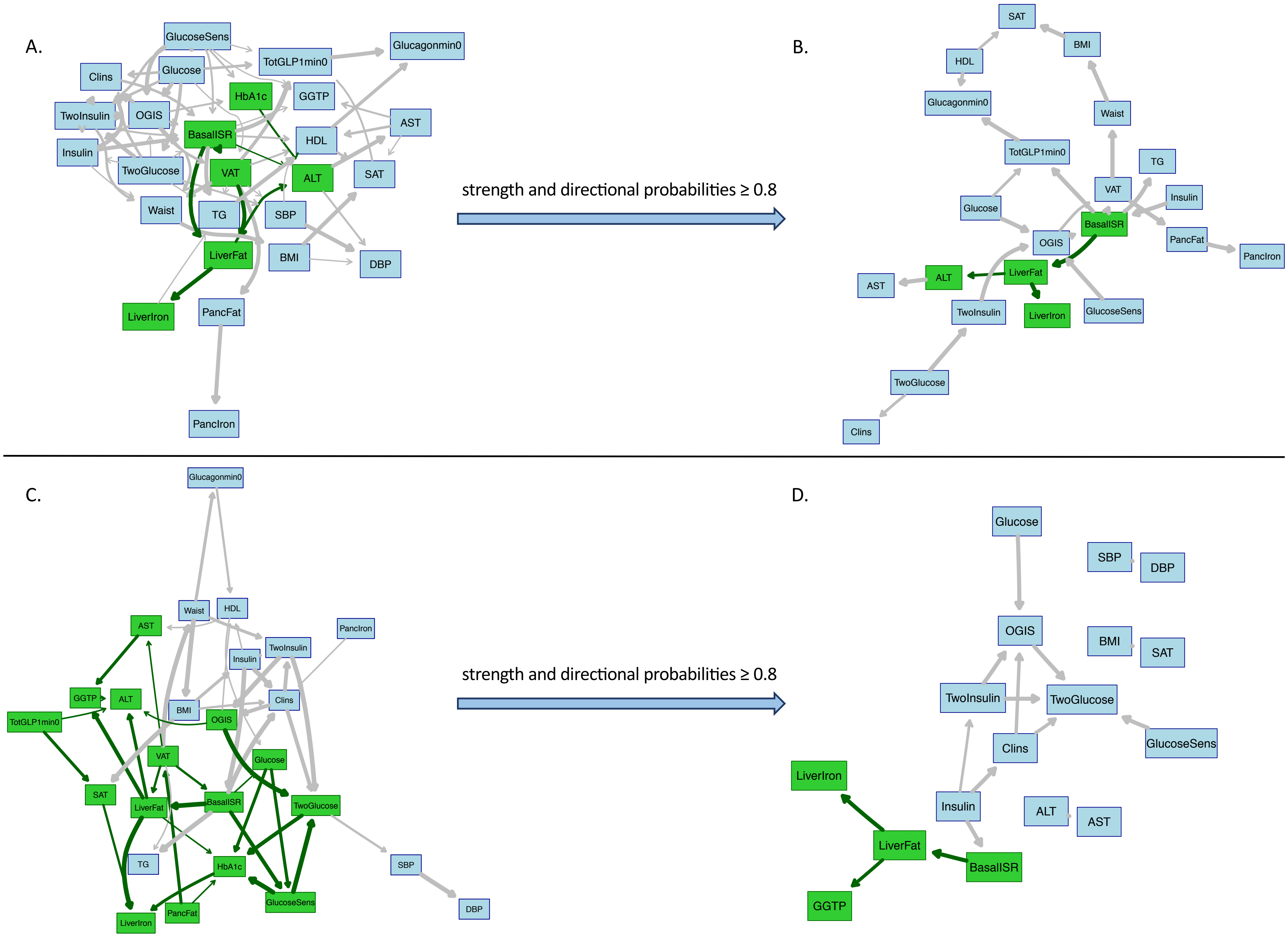

### Supplemental figure 3

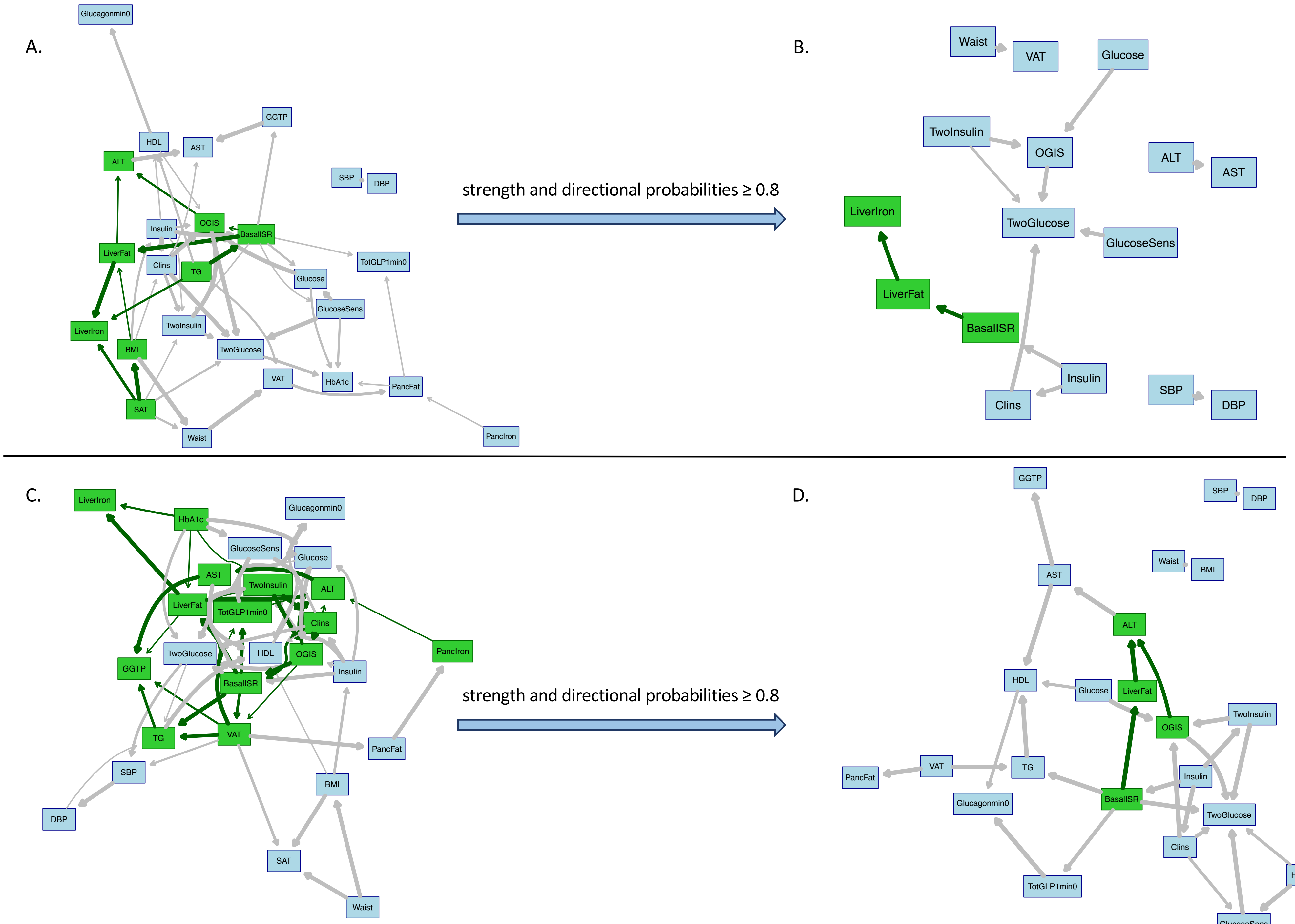

### Supplemental figure 4

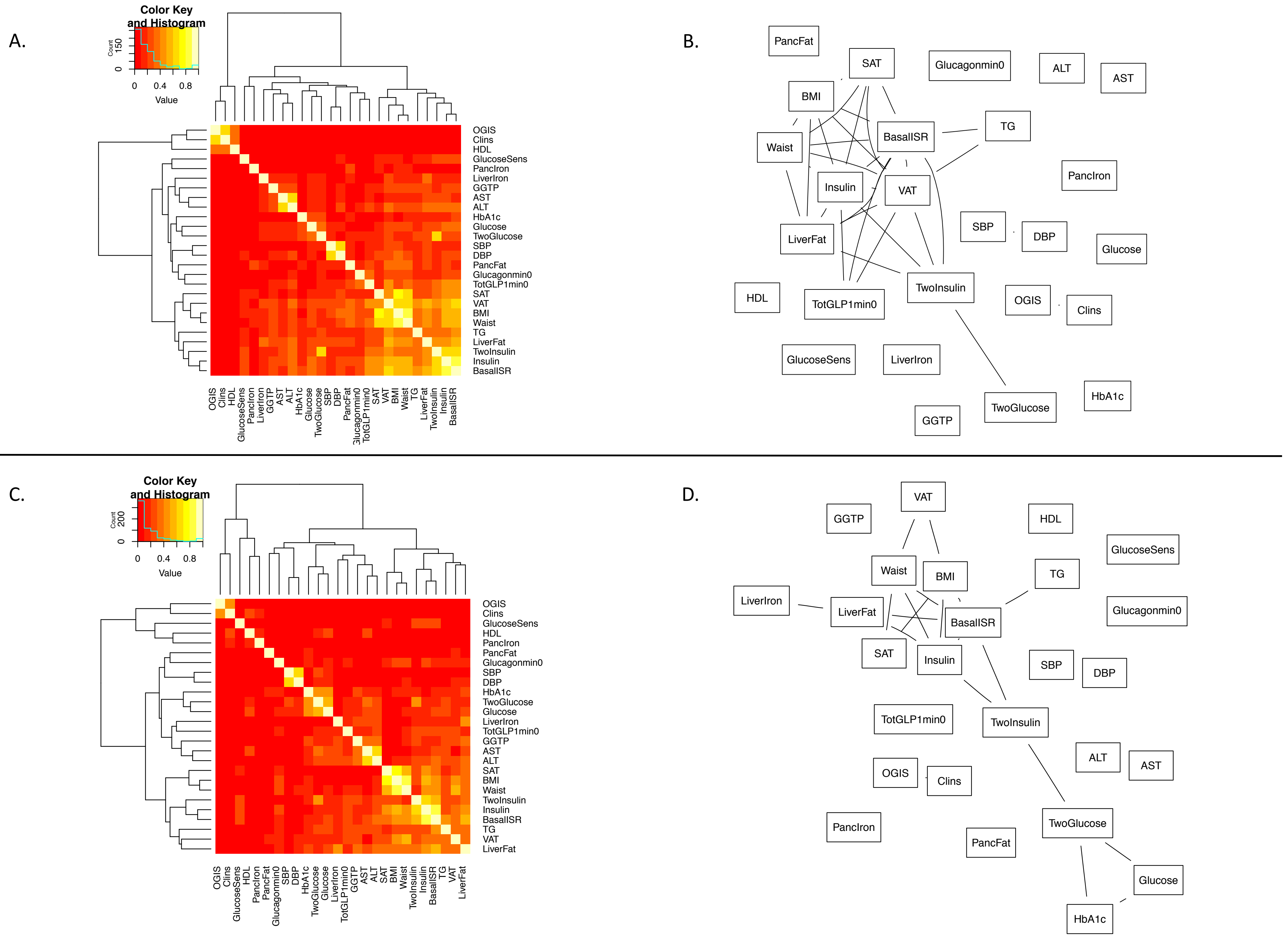

### Supplemental figure 5

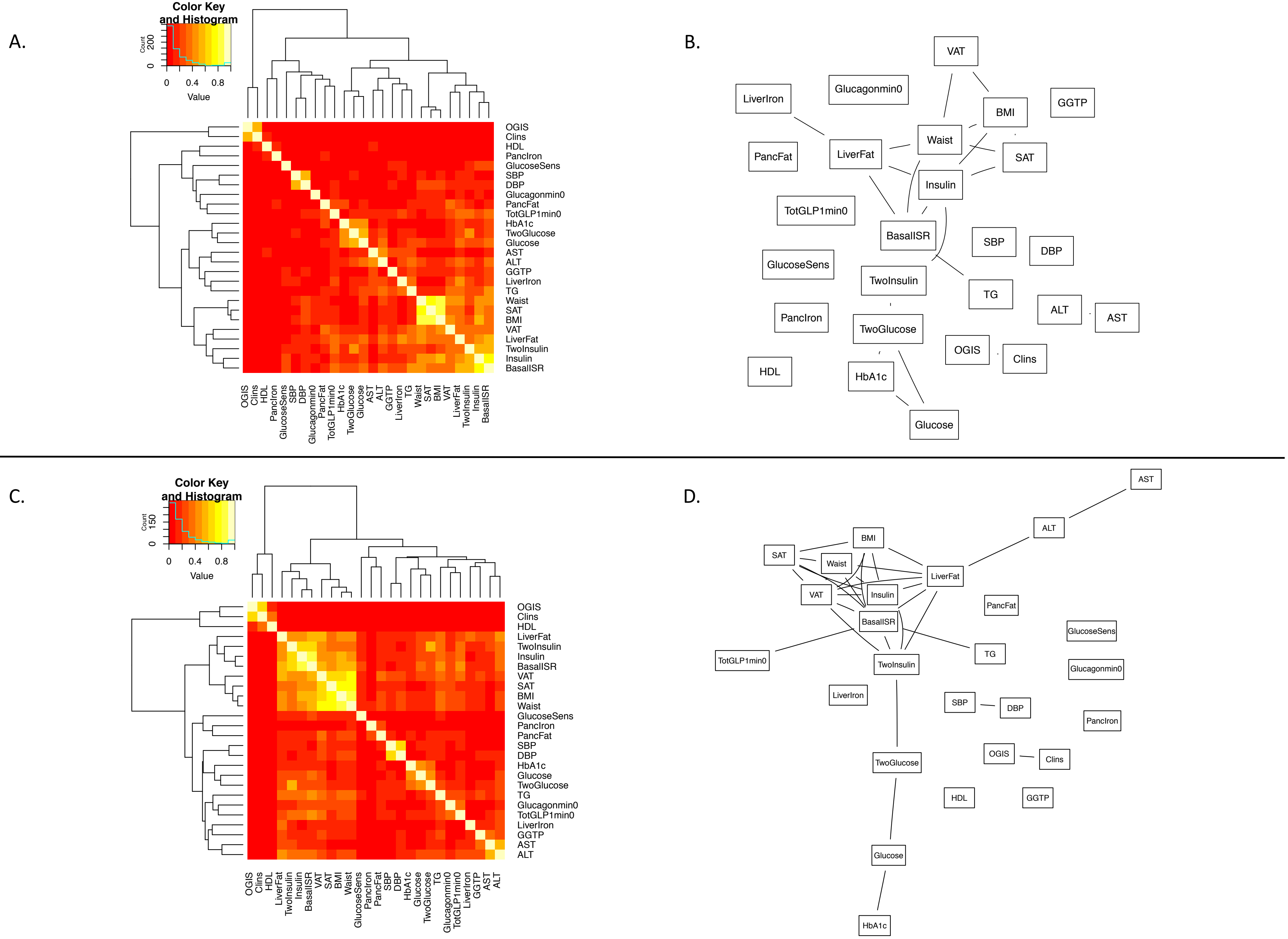

### Supplemental figure 6

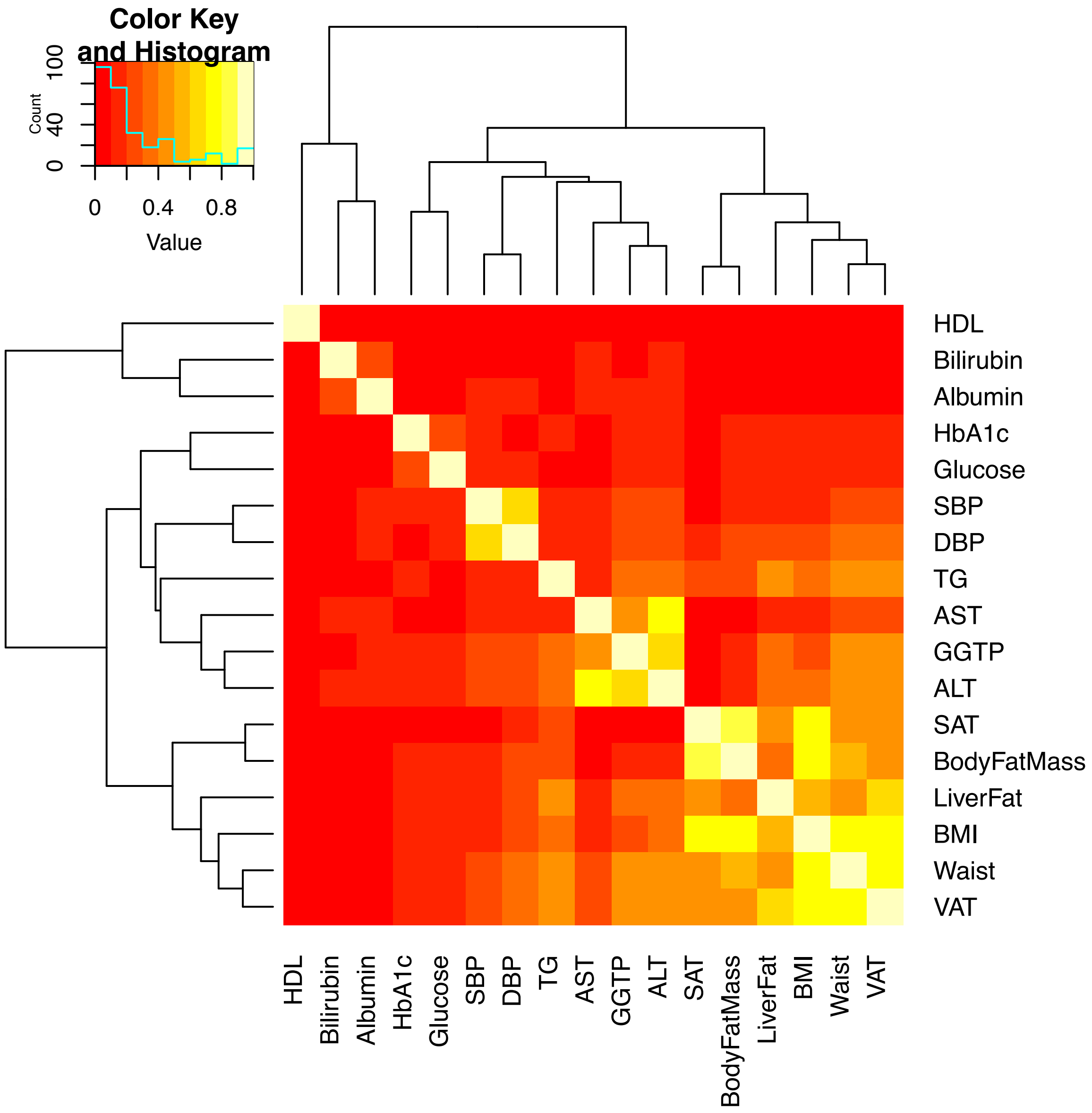

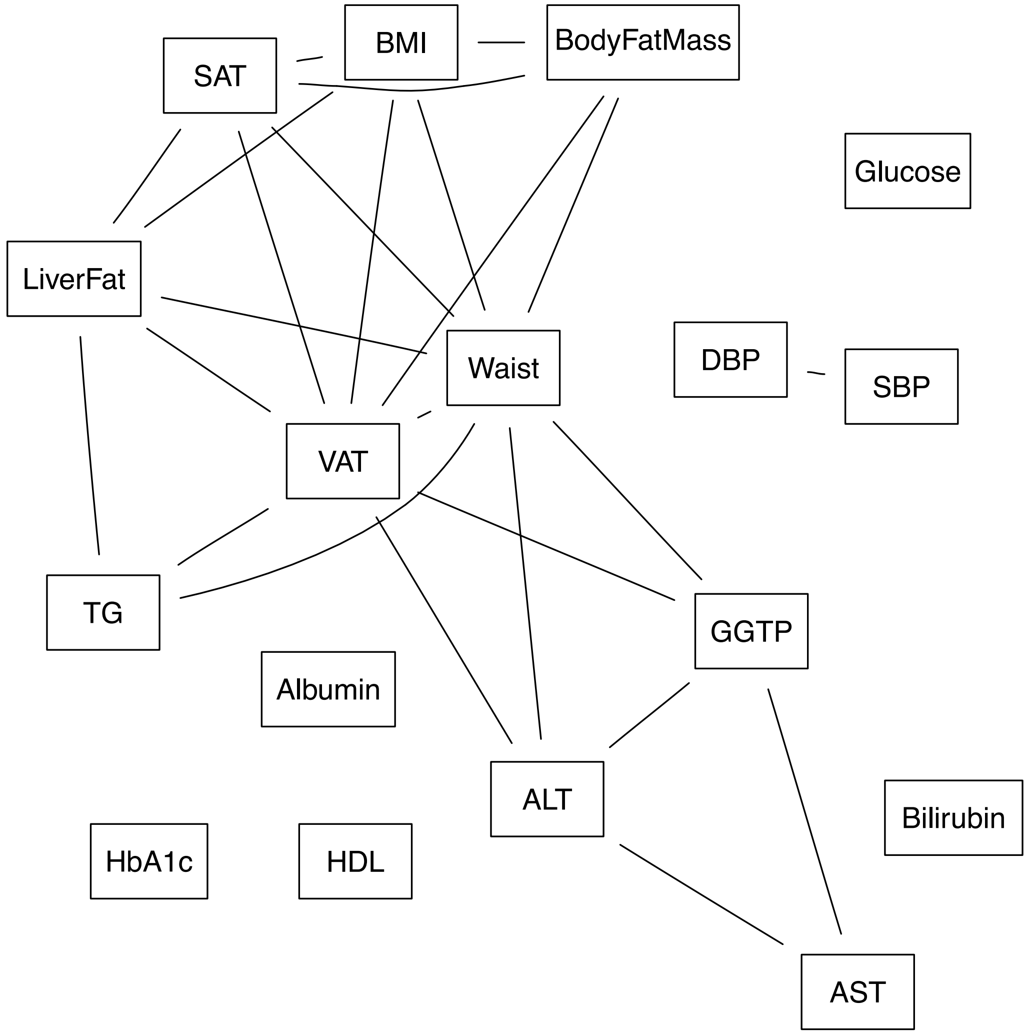

### Supplemental figure 7

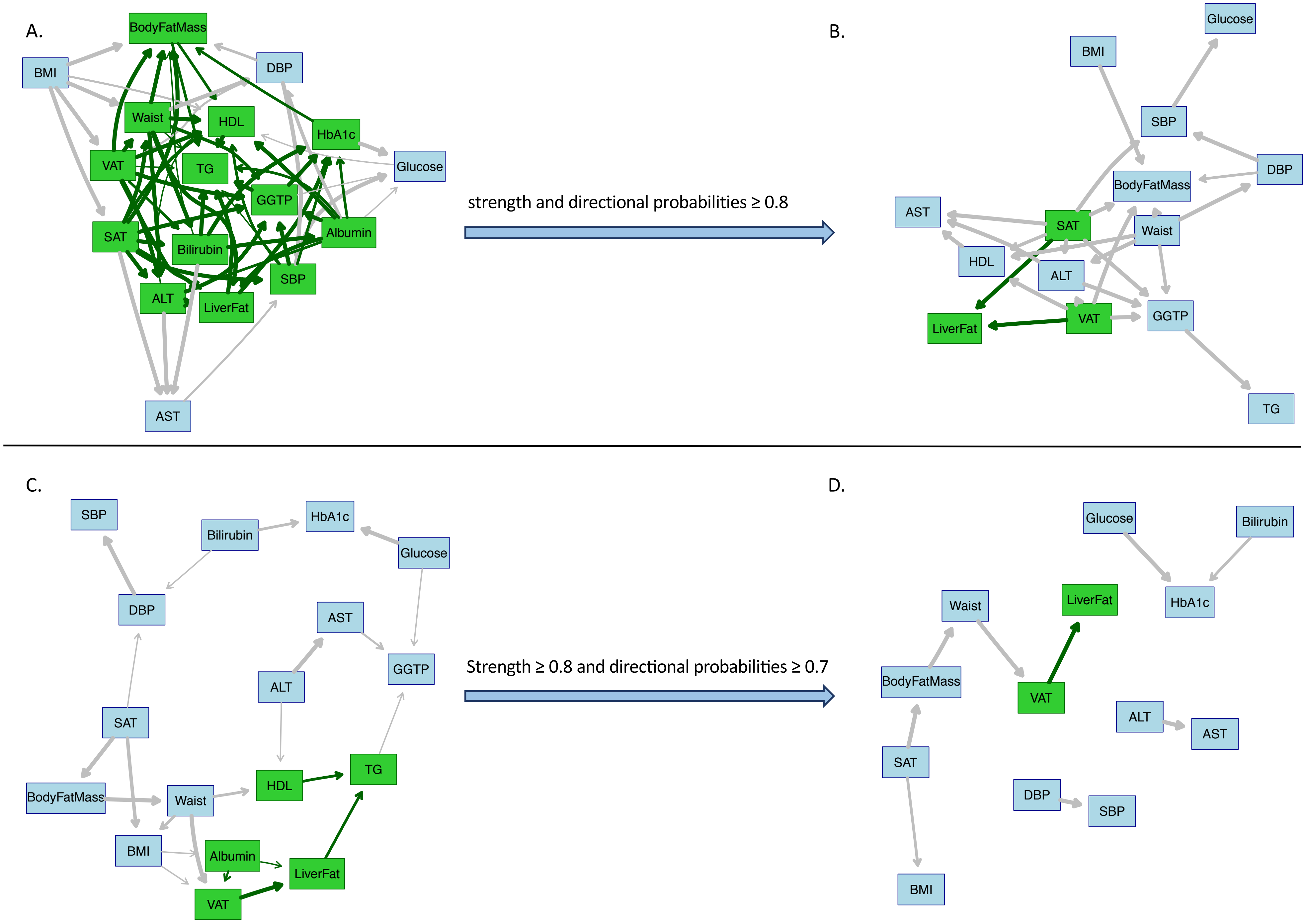

### Supplemental figure 8

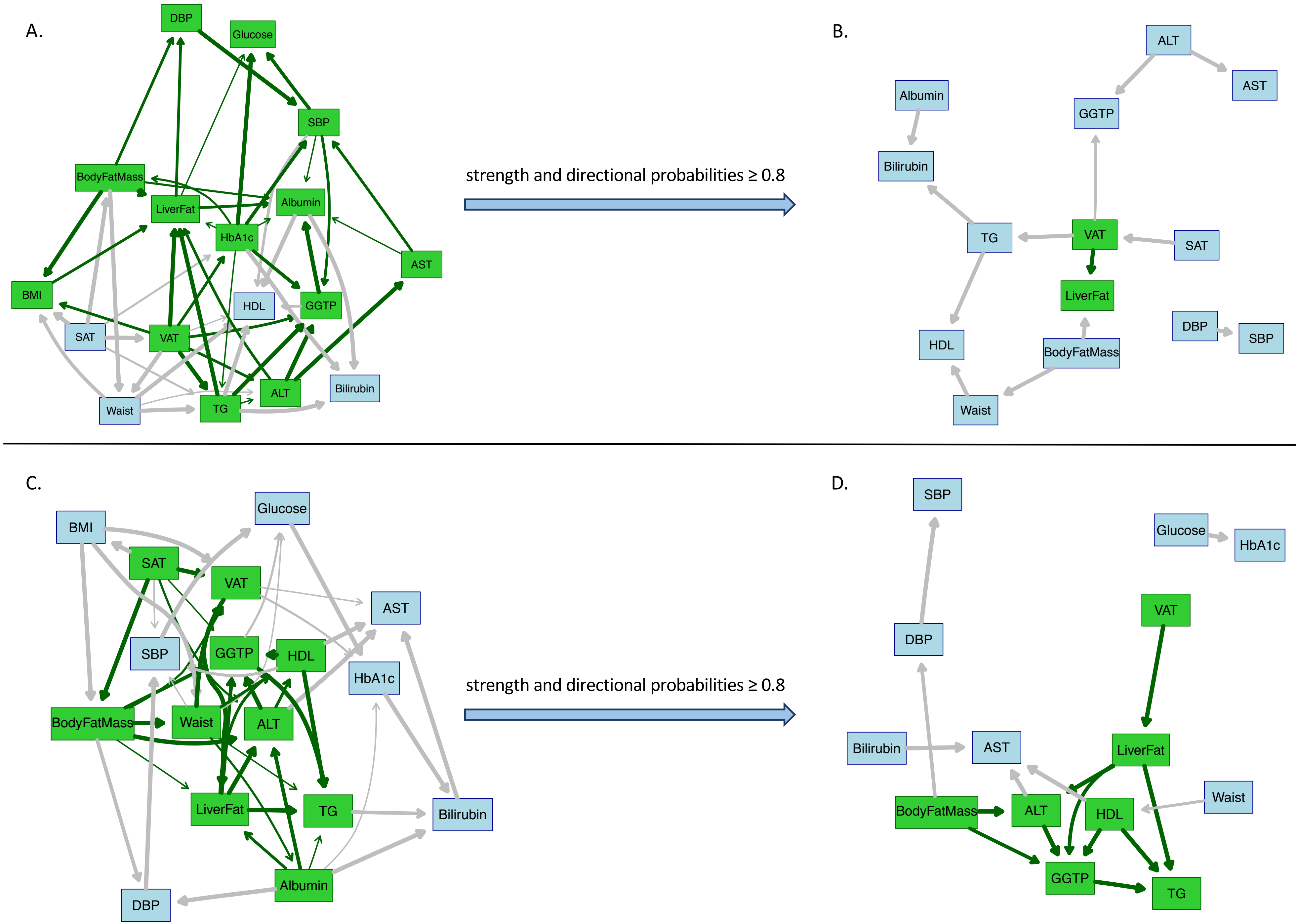

### Supplemental figure 9

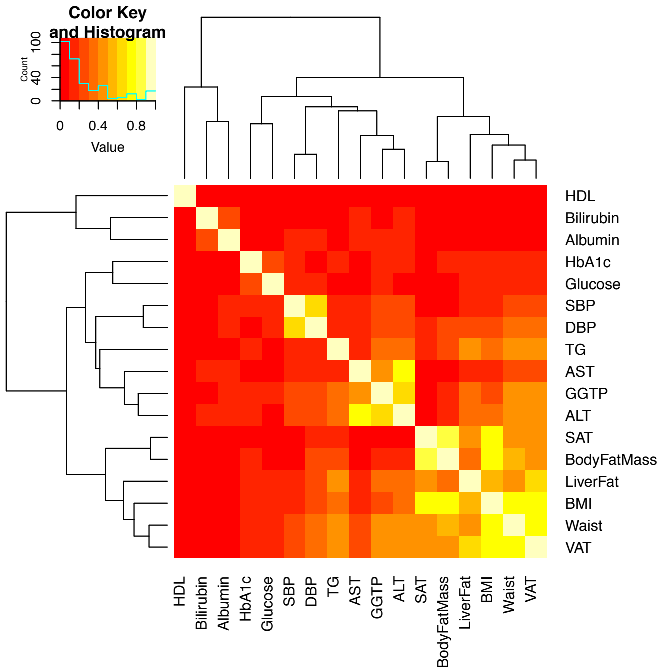

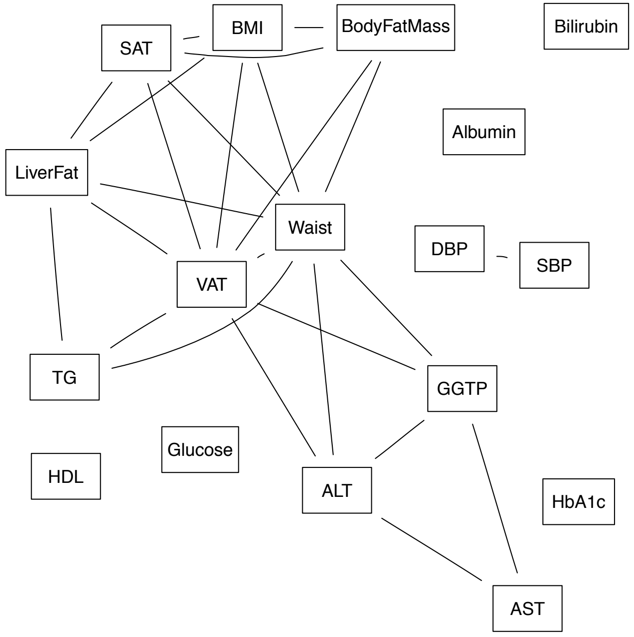

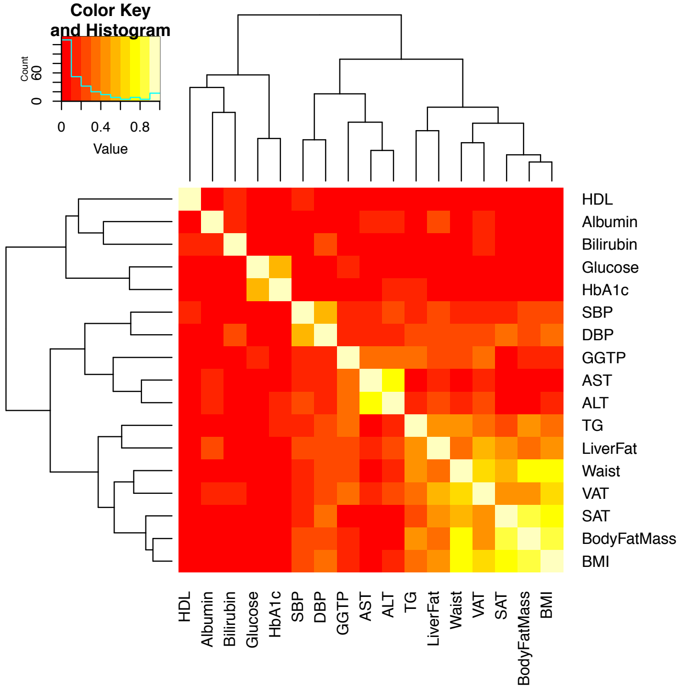

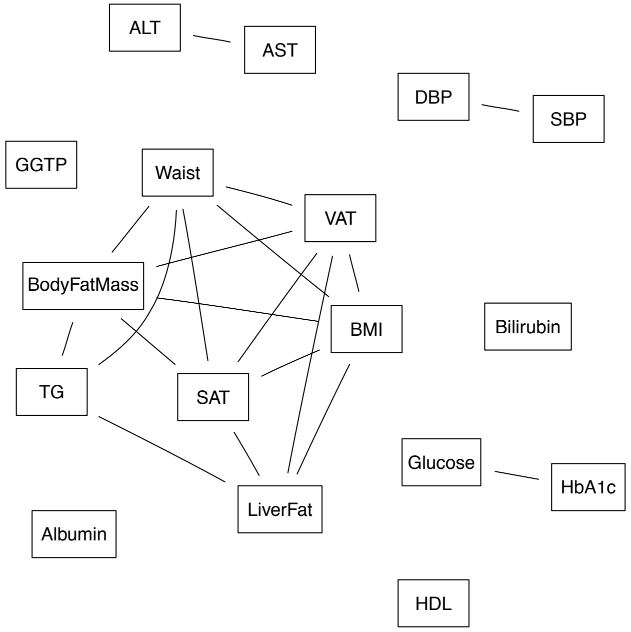

### Supplemental figure 10

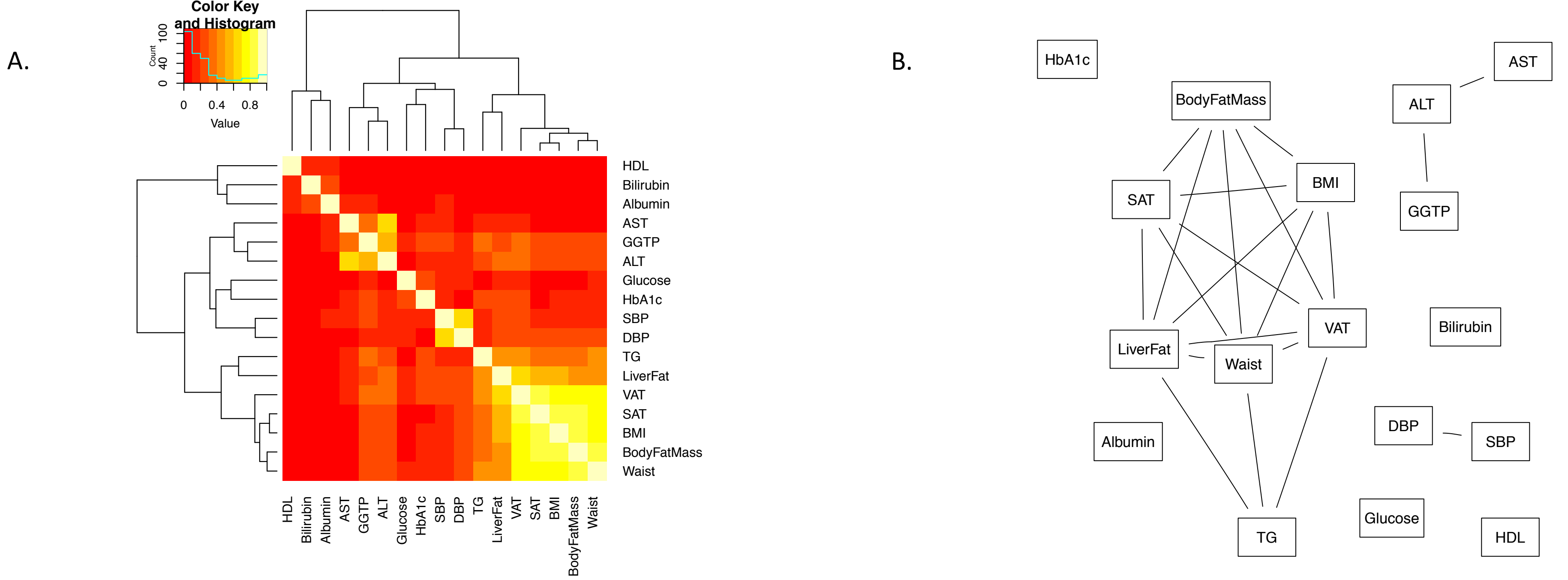

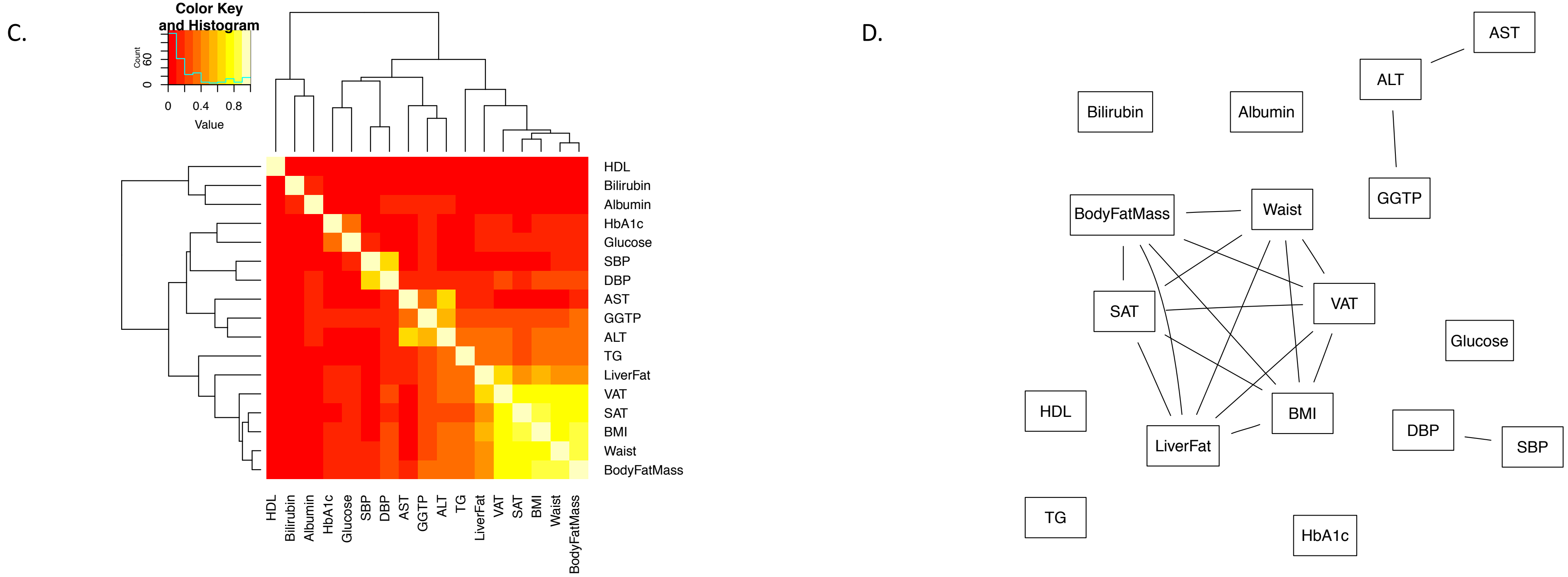

### Supplemental figure 11

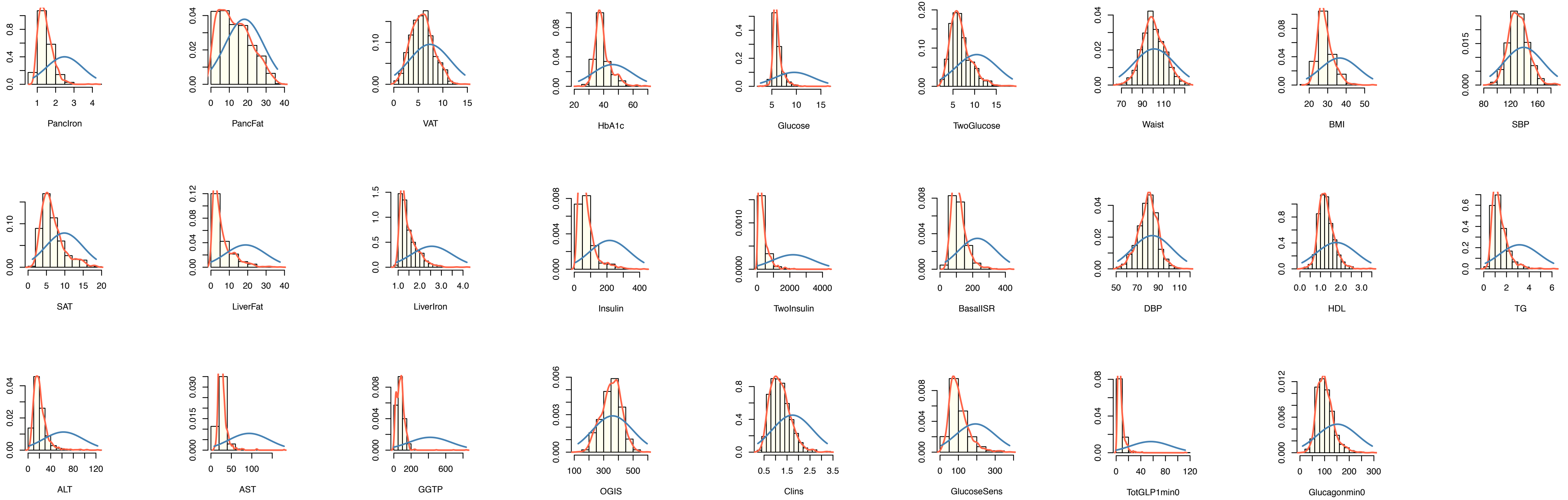

### Supplemental figure 12

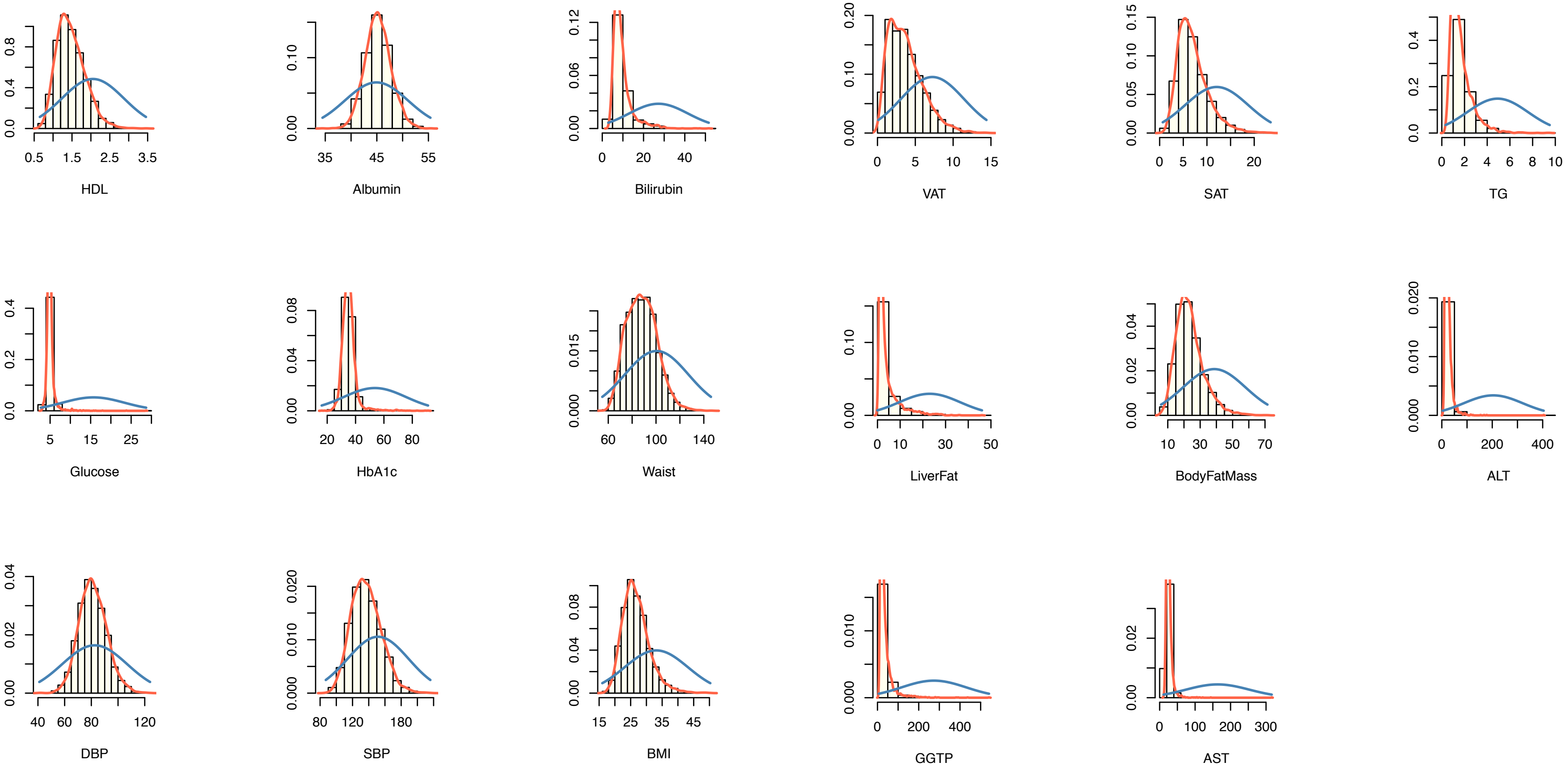
